# Stochastic modelling of early-stage COVID-19 epidemic dynamics in rural communities in the United States

**DOI:** 10.1101/2024.10.11.24315297

**Authors:** Punya Alahakoon, Peter G. Taylor, James M. McCaw

## Abstract

COVID-19, caused by the severe acute respiratory syndrome coronavirus 2 (SARS-CoV-2), has affected millions of people around the globe. We studied the spread of SARS-CoV-2 across six rural counties in North and South Dakota in the United States. The study period was from early March 2020 to mid-June 2021, during which non-pharmaceutical interventions (NPIs) were in place. The end of the study period coincided with the emergence of the Delta variant in the United States. We modelled the transmission dynamics in each county using a stochastic compartmental model and analysed the data within a Bayesian hierarchical statistical framework. We estimated key epidemiological and surveillance parameters including the reproduction number and reporting probability. We conducted a series of counterfactual analyses in which NPIs were lifted earlier and by varying degrees, modelled as an increase in the transmission rate. Under this range of plausible alternative responses, increases in case counts varied from negligible to substantial, underscoring the importance of timely public health measures and compliance with them. From a methodological perspective, our study demonstrates that despite the inherent high variability in epidemic behaviour in small rural communities, the combination of stochastic modelling and application of Bayesian hierarchical analyses enables the estimation of key epidemiological and surveillance parameters and consideration of the potential impact of alternative public health measures in small low population density communities.

## 1 Introduction

The impact of early-stage COVID-19, caused by the severe acute respiratory syndrome coronavirus 2 (SARS-CoV-2), was far from uniform across the world. This impact was notably disproportionate across different regions within the United States (Kettl, 2020; Lurie & Sharfstein, 2023).

Evidence shows that transmission dynamics were affected by factors such as urbanization, population density, and population age (Delamater, Street, Leslie, Yang, & Jacobsen, 2019; Pandey, Gu, & Ramaswami, 2022; Sy, White, & Nichols, 2021). Furthermore, in the United States, the per capita burden of disease and the relative burden on the healthcare system due to COVID-19 was high in rural areas compared to urban areas (Miller, Becker, Grenfell, & Metcalf, 2020; Sy et al., 2021). During the early stages of the pandemic, it was observed that there was substantial difference in the uptake of non-pharmaceutical interventions between rural and urban areas in the United States and throughout the world (Ejigu et al., 2021; Haischer et al., 2020).

Although many modelling studies have focused on understanding the transmission dynamics and impact of SARS-CoV-2 in high-density populations (Bo et al., 2021; Kissler, Tedijanto, Goldstein, Grad, & Lipsitch, 2020; Kucharski et al., 2020), less attention has been paid to rural communities (Mueller et al., 2021). Inspired by a news article in the *New York Times* on this very topic (Leatherby, 2020), we studied SARS-CoV-2 transmission dynamics in six rural counties in North and South Dakota.

We considered infections that occurred in the rural counties Miner, Buffalo, and Faulk in South Dakota (SD) and Towner, Eddy, and Golden Valley in North Dakota (ND) during the period prior to the *Delta* variant gaining dominance in the United States (US); that is, from early March 2020 to mid-June 2021. According to a study by the Centers for Disease Control and Prevention (CDC) (Paul et al., 2021), during April 11–24, 2021, the *Alpha* variant represented 66% of US infections, and 5% of infections were from the *Gamma* variant. During this early phase of the pandemic, vaccines were not yet available. Disease transmission was mitigated through non-pharmaceutical interventions such as social distancing measures, mask-wearing, and limiting mobility by restricting public transport availability, event cancellations, school closures, working from home requirements and stay-at-home orders (Banholzer et al., 2021; Liu et al., 2021; Reiner et al., 2021).

The main objectives of this study were two-fold. Firstly, we aimed to illustrate the value of hierarchical statistical methods for analysing epidemic dynamics in small communities. Accordingly, we modelled the epidemics using a stochastic model. To parameterise the model, we adopted a Bayesian hierarchical approach that enabled simultaneous modelling of multiple counties. Through this framework, we obtained estimates for both hyper-parameters (describing the distribution from which county level parameter estimates are drawn) and the county-specific estimates themselves.

Secondly, after conducting the hierarchical analysis and estimating the epidemiological parameters for these counties, we aimed to explore the potential impact on COVID-19 infections of alternative implementations of non-pharmaceutical interventions (NPIs). We quantified this impact through a counterfactual analysis, examining what might have occurred had certain public health measures been relaxed earlier or had different impacts on transmission.

## 2 Materials and methods

### 2.1 Data

We sourced publicly available datasets from the COVID-10 Data Hub (Guidotti & Ardia, 2020). These datasets contained daily COVID-19 case numbers, deaths, the presence and magnitude of the public health intervention measures, and the daily vaccine intake from mid-December 2020. We studied three rural counties each from South and North Dakota, all of which had populations of fewer than 2500 people in 2019.

We selected these six counties because each was sufficiently distant from the others so that it was reasonable to assume that the disease dynamics in each of the selected counties evolved independently from the others (see Figure 1). See Supplementary Material S1 for a discussion of the available data and its pre-processing.

**Figure 1:**
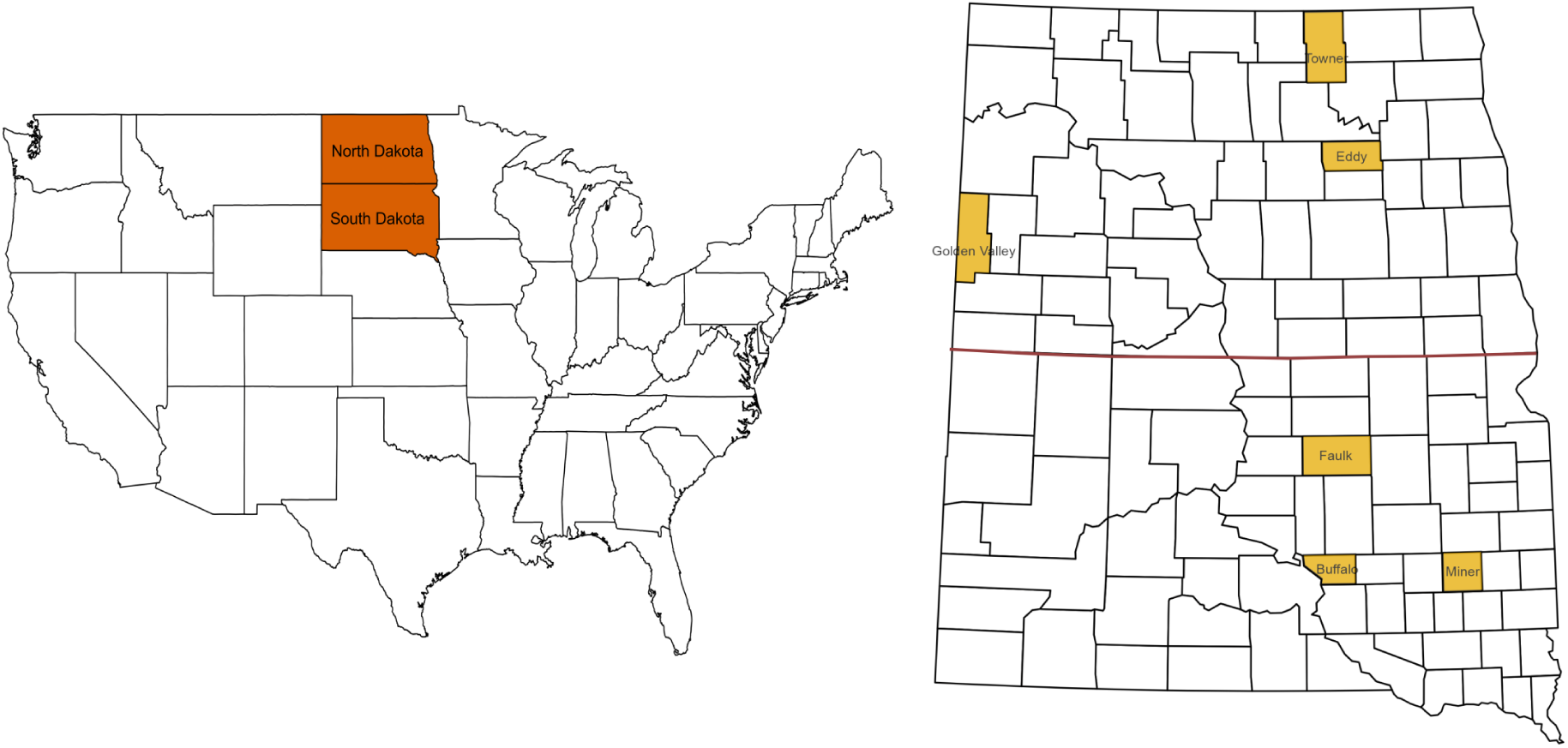
**Left:** Map of the United States with South and North Dakota in brown. **Right:** The South and North Dakota counties used in this study are in yellow.

Figure 2 depicts the daily infection trends and vaccine uptake from early March 2020 to mid-June 2021. Additionally, Table 1 summarises the six counties under study. We obtained values for the population sizes of these counties in 2019 from various sources, as detailed in Supplementary Material S1, which also includes a graphical representation of the counties’ intervention measures.

**Figure 2:**
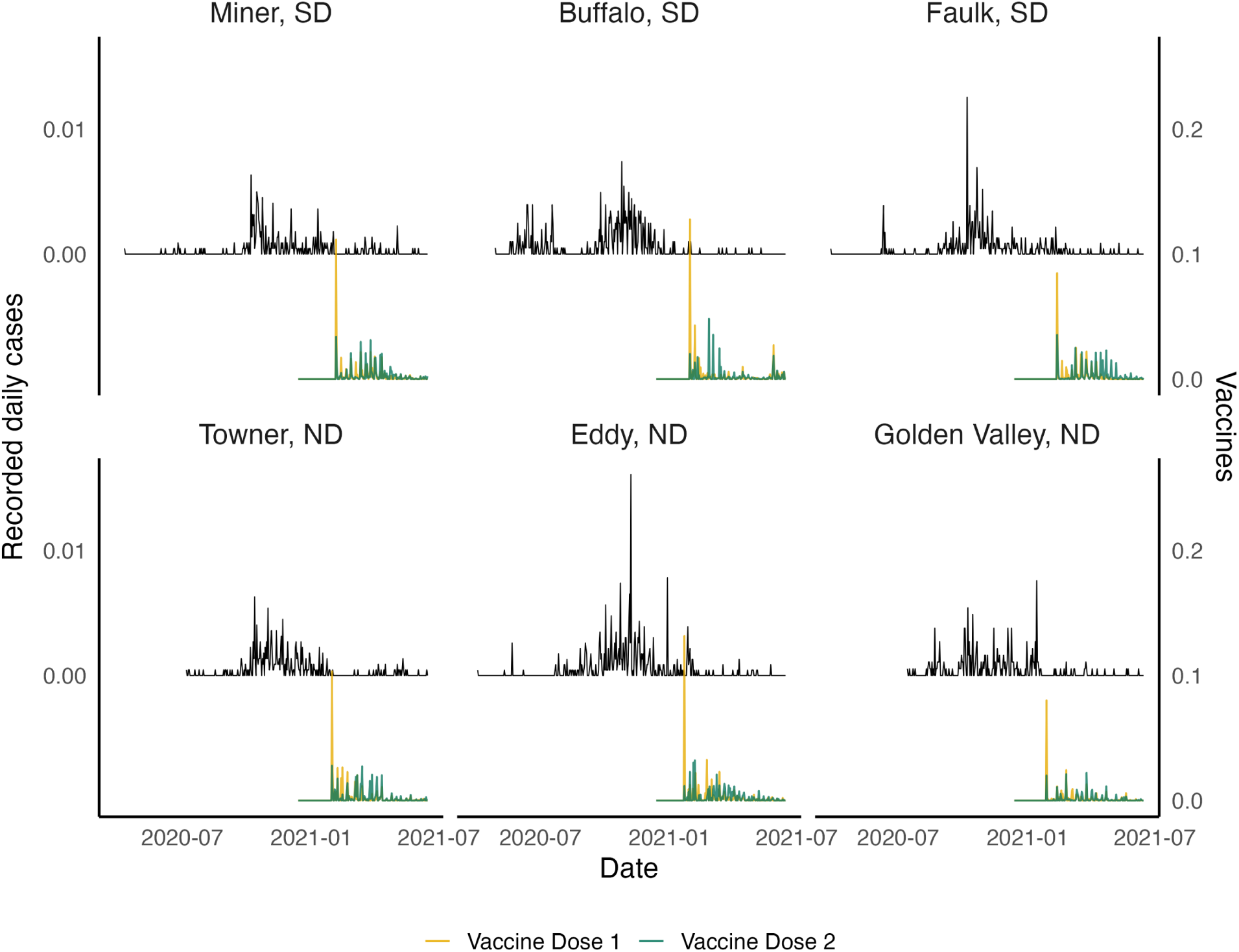
The number of cases and vaccinations recorded each day (expressed per capita) in Miner, Buffalo and Faulk counties in South Dakota and Towner, Eddy, and Golden Valley counties in North Dakota.

**Table 1:** Summary of the details of COVID-19 cases from early March 2020 to mid-June 2021

| County | Population size in 2019 | Total number of infections | Total number of deaths due to COVID-19 | Total number of first doses of vaccine | Total number of second doses of vaccine |
| --- | --- | --- | --- | --- | --- |
| Miner, SD | 2211 | 308 | 10 | 913 | 882 |
| Buffalo, SD | 2026 | 436 | 13 | 794 | 659 |
| Faulk, SD | 2312 | 377 | 13 | 753 | 877 |
| Towner, ND | 2224 | 330 | 11 | 830 | 801 |
| Eddy, ND | 2301 | 488 | 6 | 984 | 927 |
| Golden Valley, ND | 1845 | 266 | 2 | 293 | 271 |

### 2.2 Modelling and assumptions

We modelled each county’s epidemic using the stochastic compartmental model (a continuous-time Markov chain) depicted in Figure 3. The state at time *t* is the vector *{S*(*t*) *. . . R*(*t*)*}* representing the numbers of individuals in each of the compartments. The population size *N* (*t*) is the sum of the number of individuals in these compartments. The epidemic is initiated with a single person in compartment *E*. Additional exposed persons enter the county at rate *b*. As the study period predates the availability of vaccines and there was negligible prior circulation of SARS-CoV-2, we assume that all remaining individuals are initially fully susceptible, that is *S*(0) = *N* (0) *−* 1.

**Figure 3:**
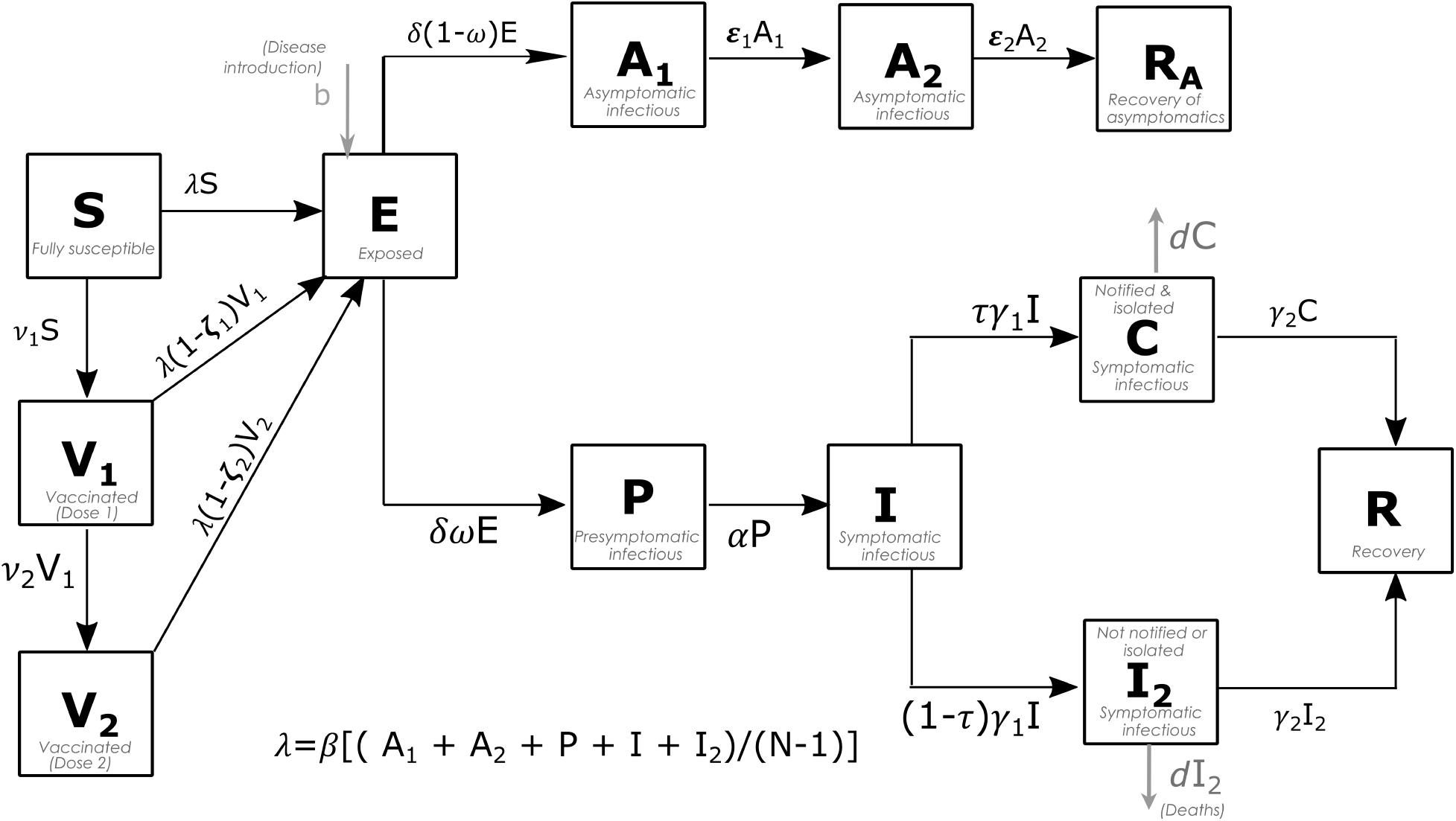
Our stochastic compartmental model for the epidemics in the US rural counties. Here, a fully susceptible (*S*) person can become exposed (*E*) to SARS-CoV-2. They can either become asymptomatically infectious (*A*_1_*, A*_2_) and recover (*R_A_*), or become pre-symptomatically infectious (*P*). A pre-symptomatically infectious person transitions to the symptomatically infectious stage (*I*). If they get tested and are notified to the health system, they are isolated (*C*). Otherwise, they transition to an addtional symptomatically infectious stage (*I*_2_). Symptomatic infections (via *C* or *I*_2_) recover (*R*). Those in compartments *C* and *I*_2_ may die due to infection with SARS-CoV-2. Vaccinated individuals (*V*_1_*, V*_2_) have reduced susceptibility to infection, but once exposed, follow the same stages of infection as a fully susceptible person.

Upon exposure, those in *S* can follow two paths. They either become asymptomatically infectious (following a path through *A*_1_ and *A*_2_) or pre-symptomatically infectious (moving to *P*). We use two asymptotically infectious compartments to ensure that the mean times spent on each branch of the model are comparable. An asymptomatic person will eventually recover (moving to *R_A_*).

A pre-symptomatically infectious person will transition to be symptomatically infectious (moving to *I*). Once an infectious person starts to show symptoms, they will either get tested and be notified to health authorities (moving to *C*). Otherwise they move to compartment *I*_2_. We denote the proportion of those who are infectious and symptomatic and that get tested and notified to the health authorities by *τ*. During the early stages of the pandemic, as per public health directions, individuals with symptoms were required to get tested, and if they tested positve for COVID-19, were required to isolate. Accordingly, we assume that all individuals in the *C* compartment take no further part in the transmission process. A person in either *C* or *I*_2_ dies with rate *d* or else eventually recovers (moving to *R*). Given our focus on the early phase of the COVID-19 pandemic, before the emergence of immunity-avoiding variants of the SARS-CoV-2 virus, we do not allow for re-infection in the model.

Vaccines became available in mid-December 2020. Noting the approximate fourteen day delay between innoculation and development of protective immunity, we incorporate their effects into the model from early January 2021. For mathematical convenience (and to maintain the Markov property of the model), we model delivery of first doses of the vaccine as a flow from *S* to *V*_1_ at rate *ν*_1_*S*. Second doses are modelled as a flow from *V*_1_ to *V*_2_ at rate *ν*_2_*V*_1_. Those vaccinated have a reduced susceptibility to infection, parameterised by *η*_1_ and *η*_2_, the proportional reduction in the probability of infection given exposure. Those infected from either *V*_1_ or *V*_2_ follow the same transition processes as described above.

We model the force of infection (*λ*) as proportional to the unweighted sum of all contagious classes (*A*_1_, *A*_2_, *P*, *I* and *I*_2_), as there is limited if any evidence for differences in the transmissibility from presymptomatic, asymptomatic and symptomatic people (Mugglestone et al., 2022).

All parameters and other related assumptions are described in Supplementary Material S2. Note that we have not accounted for age structure. Doing so would dramatically increase the number of parameters required to be estimated, adding to the complexity of the model. Particularly given the limitations of the data, we expect this would result in unreliable parameter estimates, due to both identifiability and computational (convergence) issues. Furthermore, our study is concerned with transmission of the SARS-CoV-2 virus, for which age effects, while still important, are (far) less dramatic than are the clinical consequences of infection with it. We also have not accounted for vaccinations that may have been delivered to those who were exposed, infectious or recovered.

### 2.3 Estimation framework

Model calibration was based on multiple data for each county, including the observed time-series of the number of daily cases, the total number of deaths during the analysis period, and the total number of first-dose and second-dose vaccinations delivered. Full details are provided in Supplementary Material S3.

We model the observed time-series data of the number of daily cases (that is, those tested and notified) as the cumulative incidence:

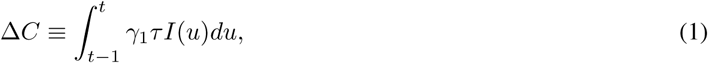

over the preceding day of the flux into *C*.

The cumulative number of first-dose vaccinations is given by ∫^∞^_0_ *ν_1_ S*(*u*)*du*. Similarly, the cumulative number of second-dose vaccinations is given by ∫^∞^_0_ *ν_2_V_1_* (*u*)*du*.

For each county we need to estimate fifteen parameters. We adopted a Bayesian hierarchical approach (Alahakoon, McCaw, & Taylor, 2022; Gelman & Hill, 2006; McGlothlin & Viele, 2018; Royle & Dorazio, 2008), choosing to model two parameters of particular interest (the transmission rate *β* and the proportion of symptomatic individuals who were tested, notified, and isolated immediately (*τ*)) as being independently drawn from a common (and so estimated) multivariate truncated normal distribution described by its hyperparameters. The remaining thirteen parameters were assumed to be county-specific (that is, independently sampled for each county with no hierarchical structure). There are many other options for setting up the overall hierarchical structure for the model (with some parameters fixed across counties, some independently sampled, and some modelled under a hierarchical structure). Our modelling choice reflects that 1) the primary focus of our study is on transmission; and 2) it is likely that there are shared attributes across counties of the pathogen (for example, transmission) and people’s behaviour (for example, test seeking behaviour). Although it would have been feasible to fix some or all of the remaining parameters across counties, we chose to retain county-specific estimates. This decision enhances the flexibility of the inference algorithm (described below), allowing it to explore a broader parameter space and identify the best-fitting models for each county. Moreover, certain parameters—such as the rate of disease re-introduction (*b*) and the rates at which fully susceptible individuals receive their first and second vaccine doses (*ν*_1_, *ν*_2_)—are inherently county-specific, reflecting local variations in exposure risk and vaccine rollout dynamics.

We conducted parameter estimation using the two-step algorithm introduced by Alahakoon et al. (2022). Step 1(a) of the algorithm performs parameter estimation considering each county independently. For this step, we used the Approximate Bayesian Computation on Sequential Monte Carlo (ABC-SMC) algorithm of Toni, Welch, Strelkowa, Ipsen, and Stumpf (2009). Samples were initially drawn from the prior distribution and then propagated through a sequence of intermediate distributions (called generations) until they could be considered to be drawn from the target (posterior) distribution (Toni et al., 2009). Step 1(b) uses the county-independent posterior samples from step 1(a) to construct an estimator for the likelihood at the hyper-parametric level, and a standard MCMC algorithm to then sample from the posterior for the hyper-parameters. Specifically, we assume that (*β_k_, τ_k_*) is sampled from a Truncated Multivariate Normal (**Ψ**, Σ, **a**, **b**) distribution, where **Ψ**, Σ, **a**, and **b** are the hyper-parameters, covariance matrix, and lower and upper truncation points respectively. In step 2 of the algorithm, parameters for each of the counties under the hierarchical model are sampled using an ABC algorithm, drawing from the posterior distribution for the hyper-parameters (constructed in step 1(b)). That is, we sampled (*β_k_, τ_k_*) from the Truncated Multivariate Normal (**Ψ**, Σ, **a**, **b**), given the estimated hyper-parameters. All other parameters (thirteen of them) were sampled by perturbing (similar to the algorithm of (Toni et al., 2009) using the same perturbation kernel) from the last generation of the marginal posterior distribution from step 1(a). Full details of the calibration of this algorithm can be found in Supplementary Material S3.

### 2.4 Policy measures implementation in South and North Dakota

As did other states in the US, South and North Dakota implemented various policy measures designed to mitigate the spread of SARS-CoV-2. These measures varied in intensity and impact. Policy measures in place during the analysis period were school closures, workplace closures, cancellation of events, stay-at-home restrictions, restricting internal and international movements, information campaigns, testing, contact tracing, facial covering mandates, and elderly people protection. Limitation on gatherings was not active most of the time in South Dakota but it was active for four to five months, on and off in North Dakota. There were no restrictions on transport in South Dakota whereas transport restrictions were recommended in North Dakota for the first three months of our analysis period. Further details and graphical illustrations regarding policy implementation in these states can be found in Supplementary Material S1.

During our analysis period, most of these interventions were at least recommended. By studying the mobility trends analysed from the CDC and Google (see Supplementary Material S7), we concluded that for all six counties, the percentage of people staying at home was significant, although it remained below 50%. There was also a noticeable reduction in travel to retail and recreation. Workplace mobility changes were negligible. While temporal variation is clearly evident in the data streams, we concluded that there was insufficient evidence to explicitly model a time-dependent transmission rate. We therefore assumed that *β* was fixed (over the time period of the study).

### 2.5 Counterfactual analyses

We were interested in investigating whether intervention measures were successful in mitigating transmission in these counties. Ideally, one would know the transmission rate before imposition of policy measures and then observe (and model) their impact. However, data we have for inclusion in the study are from a period where policy measures were already in place. While this prevented direct estimation of their impact, we sought to characterise (through parameter estimation) the epidemics occurring under the presence of NPIs in each county, and then explore through a counterfactual analysis the potential outcomes had those NPIs been been lifted earlier and by varying degrees.

We studied the impact of relaxing the interventions after two weeks, one month, and two months from the start of our analysis period. For each of these counterfactual scenarios, we studied the impact on the numbers of cases for each county if the transmission rate increased (assuming the probability of notification to health authorities given symptoms (*τ*) is unchanged).

Studies such as Kissler et al. (2020) and Ferguson et al. (2020) have demonstrated the public health utility of NPIs. In general, the stringency of (and compliance with) non-pharmaceutical interventions is reflected in mobility patterns (Gibbs et al., 2022; Golding et al., 2023; Snoeijer, Burger, Sun, Dobson, & Folarin, 2021). Most directly, mobility reflects the level of contact between members of the population, and in turn, the level of contact reflects transmission Nouvellet et al. (2021), captured by the parameter *β* in our model. Likely mediated by behavioural and social factors, provisions such as mask wearing and information campaigns also correlate broadly with mobility patterns and so are reasonably captured for our purposes by *β* (Golding et al., 2023). As described above for South and North Dakota, mobility was reduced from baseline levels by up to (approximately) 50% throughout the study period. Accordingly, we considered three levels of relaxation in the counterfactual analyses: an increase in the force of infection by 20%, 60%, and 80%. Broadly, an increase of 20% corresponds to an assumption of maintenance of relatively stringent NPIs, while an increase of 80% corresponds to an assumption of return to near-baseline levels of mobility (and so transmission).

We computed the anticipated impact for each counterfactual as follows. For each sample from the joint posterior distribution of a county, we multiplied *β* by a factor of 1.2, 1.6 and 1.8 at the different hypothesised change-point times of NPI relaxation (two weeks, one month, or two months). Under these nine scenarios, we generated sample paths from our model, with the same initial conditions for each county. Further details can be found in the Supplementary Material.

## 3 Results

Figure 4 shows the 50%, 75%, and 95% curve-wise intervals (computed using curve_interval with default settings (mhd: mean halfspace depth method) from R-package ggdist (Fraiman & Muniz, 2001; Juul, Græsbøll, Christiansen, & Lehmann, 2021; Kay, 2022)) of the smoothed 7-day moving average from re-sampled paths of the six counties, generated by re-simulating sample paths from our model using the samples from the joint posterior distribution of each county (Supplementary Material S5 shows unsmoothed re-sampled paths). These simulated paths present posterior predictive checks (Gelman et al., 2013), which assess the predictive power of our proposed model. Figure 5 illustrates the simulated peak day vs. peak size for these counties. To mitigate the impact of the observed noise, we calculated the seven-day moving averages of the cases in each county and used these to calculate the observed peak time and size. Generally, the generated paths of the cases aligned well with the observed paths.

**Figure 4:**
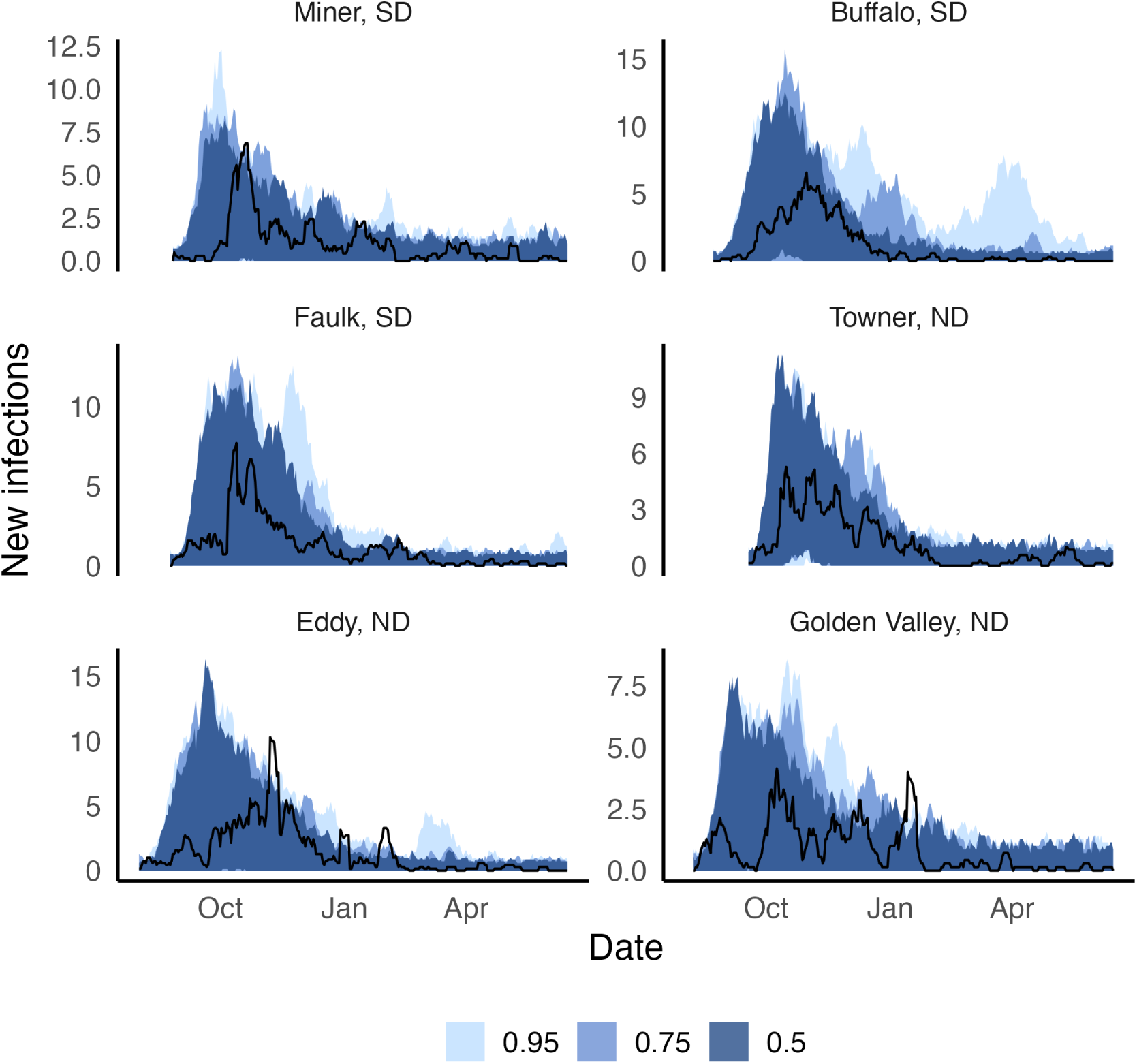
Re-sampled paths for the counties. The blue ribbons represent 50%, 75%, and 95% curve-wise intervals based on infections from 7-day moving average. The black lines show observed data, summarised as 7-day moving average.

**Figure 5:**
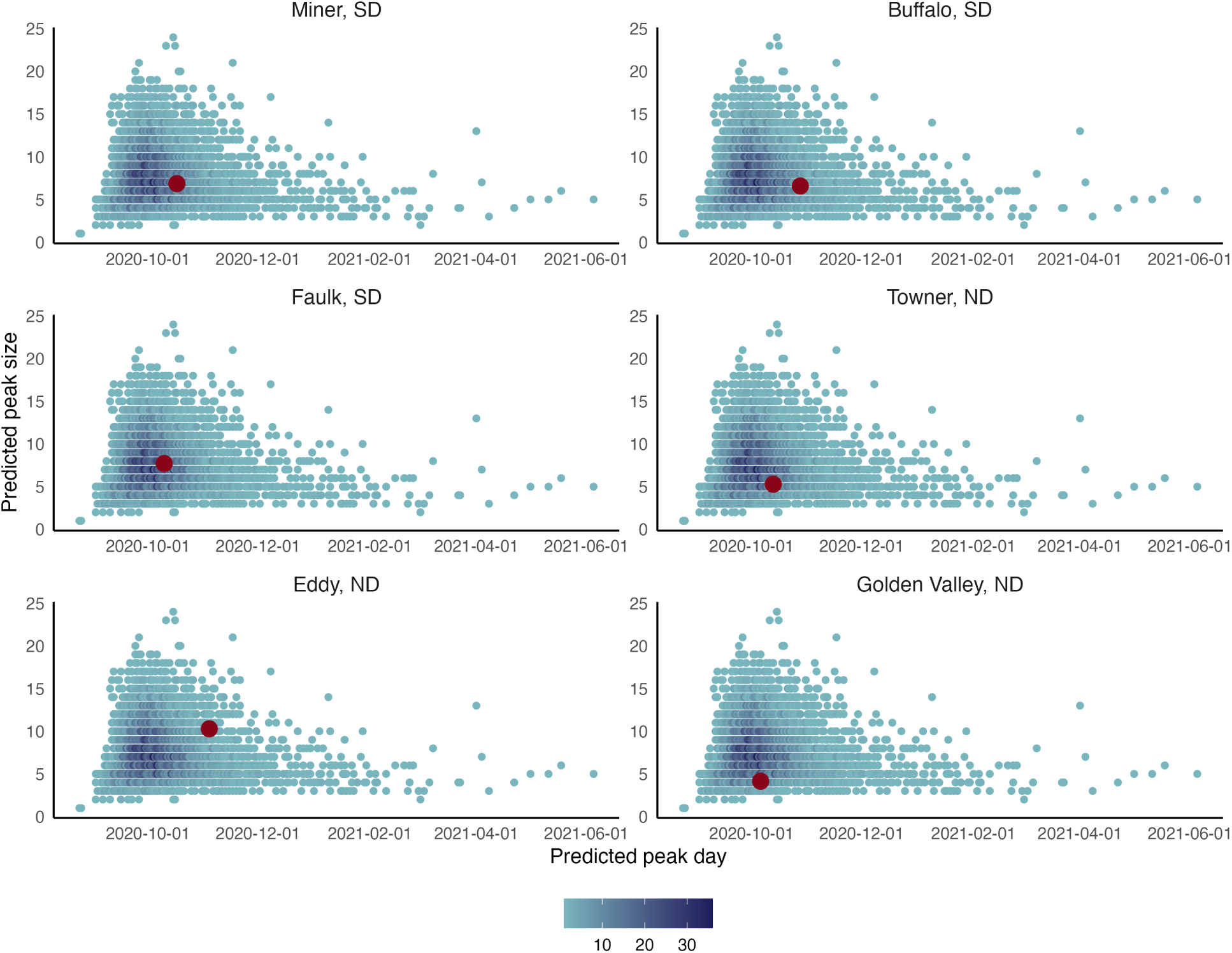
Plots of simulated peak day vs. peak size for the six counties. Observed peak days and sizes (calculated from the seven-day moving average path) are plotted in red.

For Miner, the median simulated peak size was 6 (observed peak size was 7), occurring between September and October 2020 (observed peak day: October 17, 2020). For Buffalo, the median simulated peak size was 9 (observed peak size was 7), occurring between September and November 2020 (the observed peak day: October 28, 2020). For Faulk, the median of the simulated peak size was 8 (the observed peak size was 8), occurring between September and November 2020 (the observed peak day: October 10, 2020). For Towner, the median of the simulated peak size was 8 (the observed peak size was 5), occurring between September and October 2020 (the observed peak day: October 13, 2020). For Eddy, the median of the simulated peak size was 9 (the observed size was 10), occurring between September and November 2020 (the observed peak day: November 4, 2020). For Golden Valley, the median of the simulated peak size was 5 (the observed simulated size was 4), occurring between September and November 2020 (the observed peak day: October 6, 2020). Refer to Supplementary Material S5 for other diagnostic figures, such as vaccine intake.

Figure 6 displays the posterior distributions of *τ*, the proportion of symptomatic individuals who were tested, notified and isolated immediately. Overall, *τ* for each of the counties ranged from 0.27 to 1 (highest posterior density (HPD) intervals), with a posterior median close to 0.6. Among the six counties, Eddy had the highest proportion of symptomatic individuals who were tested, notified and isolated immediately [median 0.637 and (0.320, 0.999) 95% HPD interval] while Towner had the lowest proportion [median 0.585 and (0.260, 0.968) 95% HPD interval]. Supplementary Material Table S10 presents values for the other counties with medians and 95% HPD intervals. Supplementary Material S5.1 presents posterior distributions for all other parameters. Additionally, the marginal posterior distribution for the population-level estimate (hyper-mean) for *τ* had a median of 0.606 [(0.105, 0.998) 95% HPD interval]. Supplementary Material S4 provides further details on this and other hyper-parameters.

**Figure 6:**
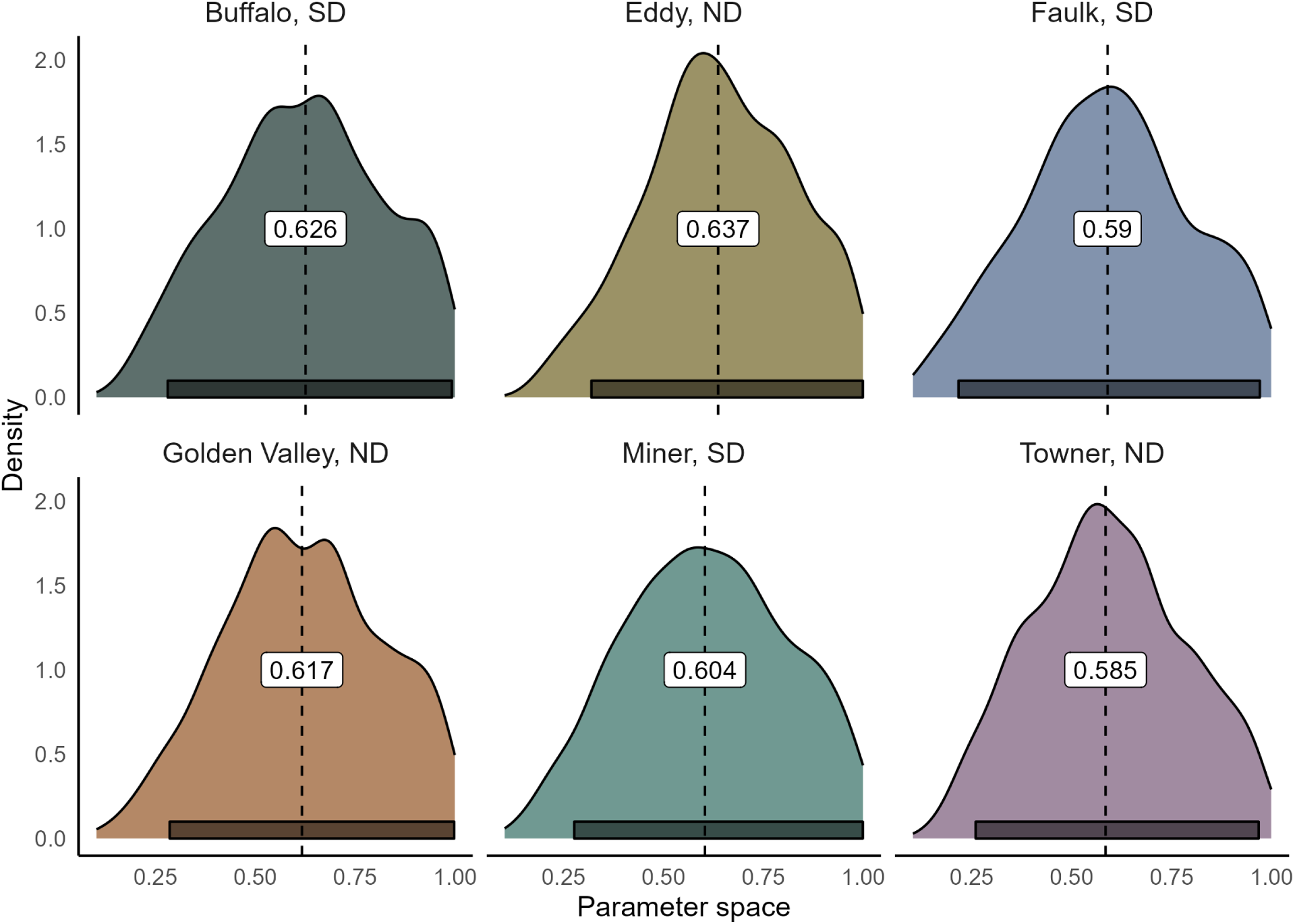
Posterior distributions of *τ*, the proportion of symptomatic individuals who were tested, notified, and isolated immediately. Each panel corresponds to one of the six counties examined in our study. The posterior median is labelled in each county and is illustrated with a black-dashed line. The 95% Highest Posterior Density (HPD) intervals are shown in grey boxes.

We calculated the basic reproduction number from the estimated parameters (see Supplementary material for details). Figure 7 illustrates the distribution of the basic reproduction number (*R*_0_) (see Supplementary Material Table S11 for the values with medians and 95% HPD intervals). Across the counties, posterior estimates for *R*_0_ ranged from just over 1 to around 1.7. Buffalo had the highest *R*_0_ [median 1.415 and (1.306, 1.527) 25% and 75% quantiles] and Miner had the lowest *R*_0_ [median 1.204 and (1.100, 1.305) 25% and 75% quantiles]. Additionally, the posterior estimate of *R*_0_ for Eddy had notably less precision in comparison to other counties.

**Figure 7:**
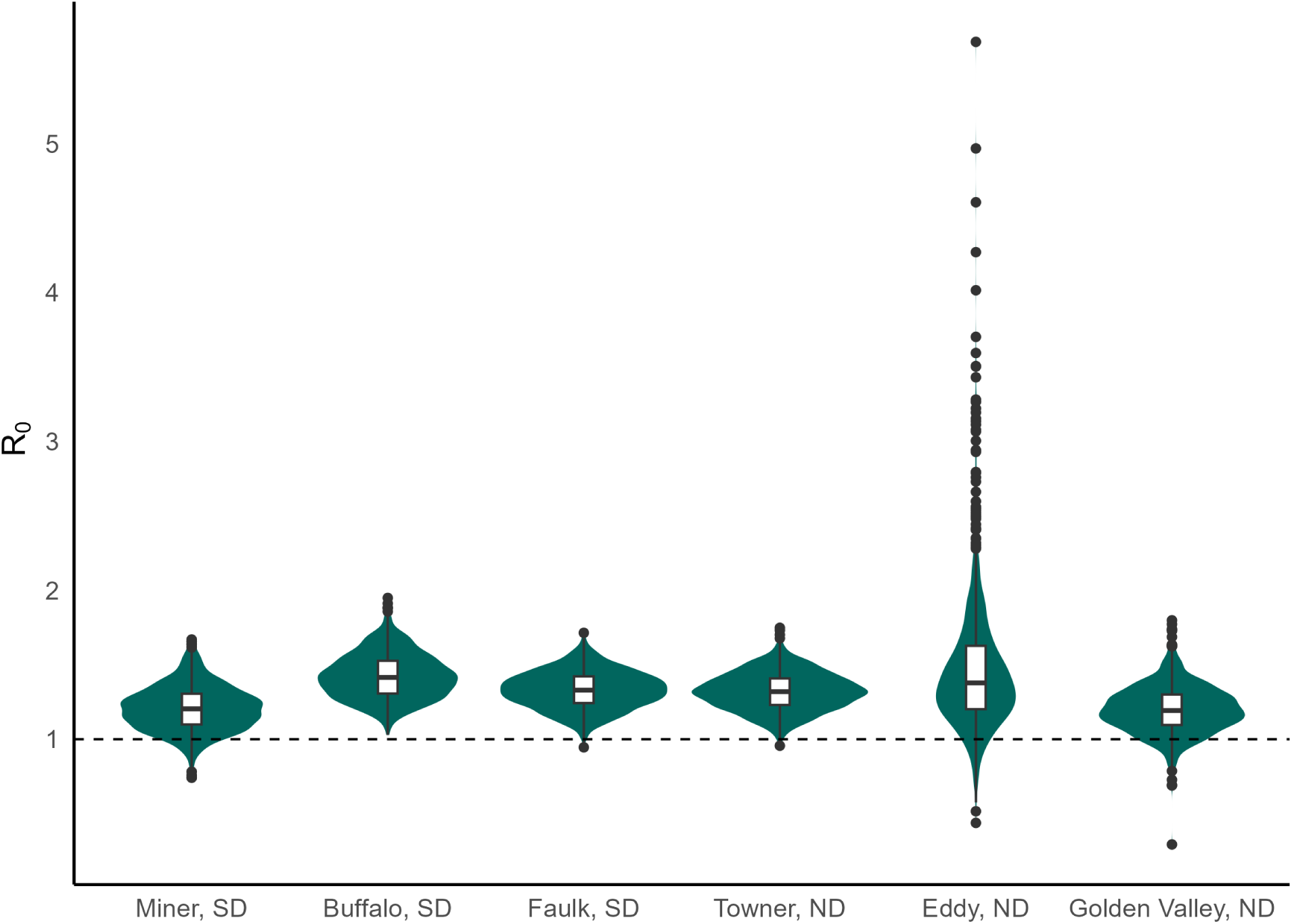
Violin plots of *R*_0_ for the rural counties

Figure 8 shows sample paths for COVID-19 cases (that is, notified infections) in Faulk SD, had the force of infection (FoI) increased by 20%, 60%, and 80% after 2 weeks, 1 month, and 2 months. Similar effects were observed in the other five counties studied. More detailed analyses for all six counties are provided in the Supplementary Material.

**Figure 8:**
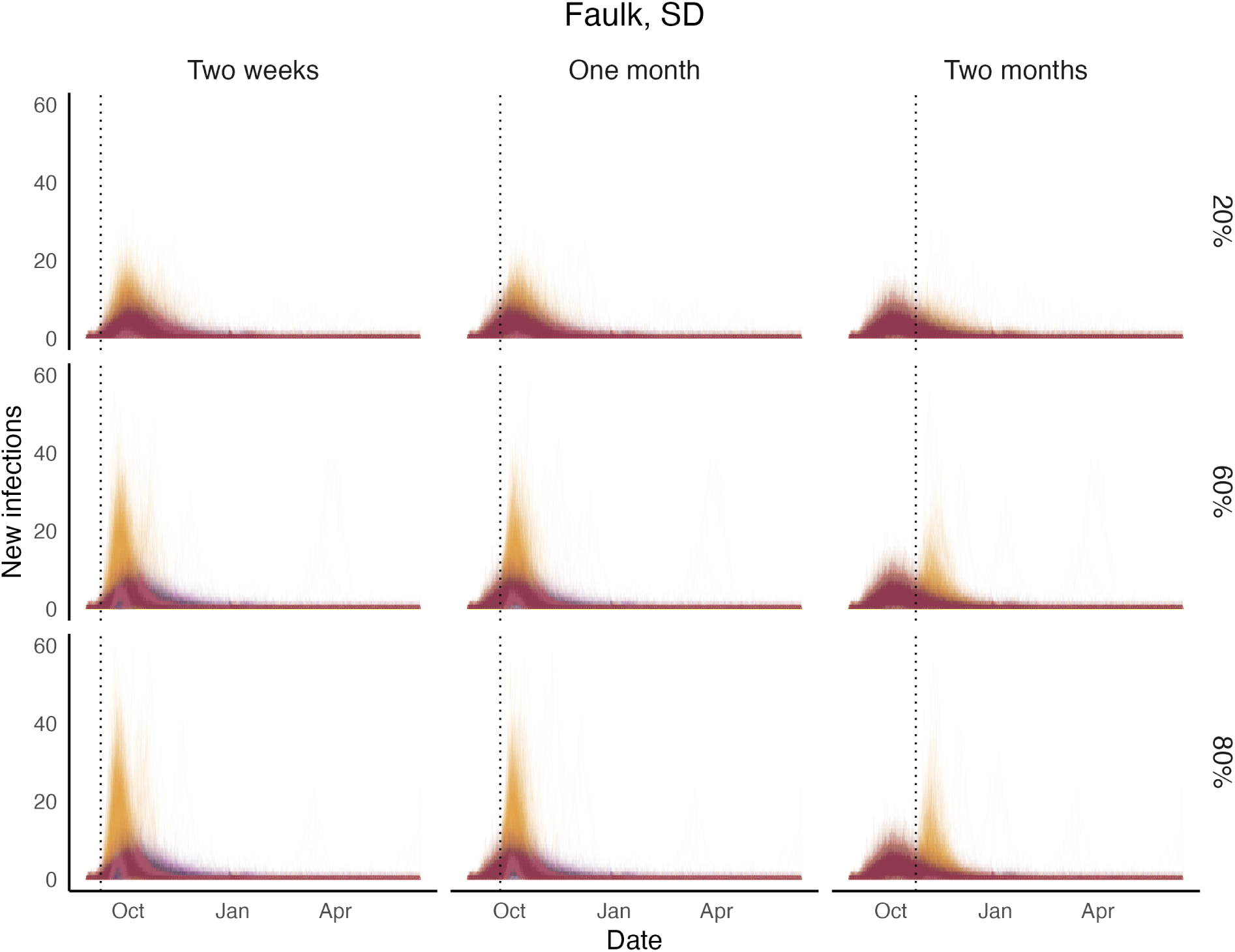
Impact of COVID-19 on Faulk, SD had the FoI increased by 20% (first row), 60% (second row), and 80% (third row) after 2 weeks (first column), one month (second column), and two months (third column). Purple paths are simulated paths with estimated FoI. Yellow sample paths are counterfactual paths.

Figure 9 presents density interval plots for the (multiplicative) change in the number of cases summed over all six counties, for different timing of relaxation and assumed increase in the FoI due to the cessation of non-pharmaceutical interventions. As expected, the greatest increase in the total number of cases occurred when the FoI was increased at the earliest point and by the largest amount we considered (80% at two weeks). This resulted in a median increase in the total number of cases across all six counties of 50.7% [(43.6%, 57.6%) 25% and 75% quantiles]. While both the timing and degree of change in the FoI are important, outcomes were more sensitive to the assumed level of increase in the FoI than to the timing of relaxation of measures.

**Figure 9:**
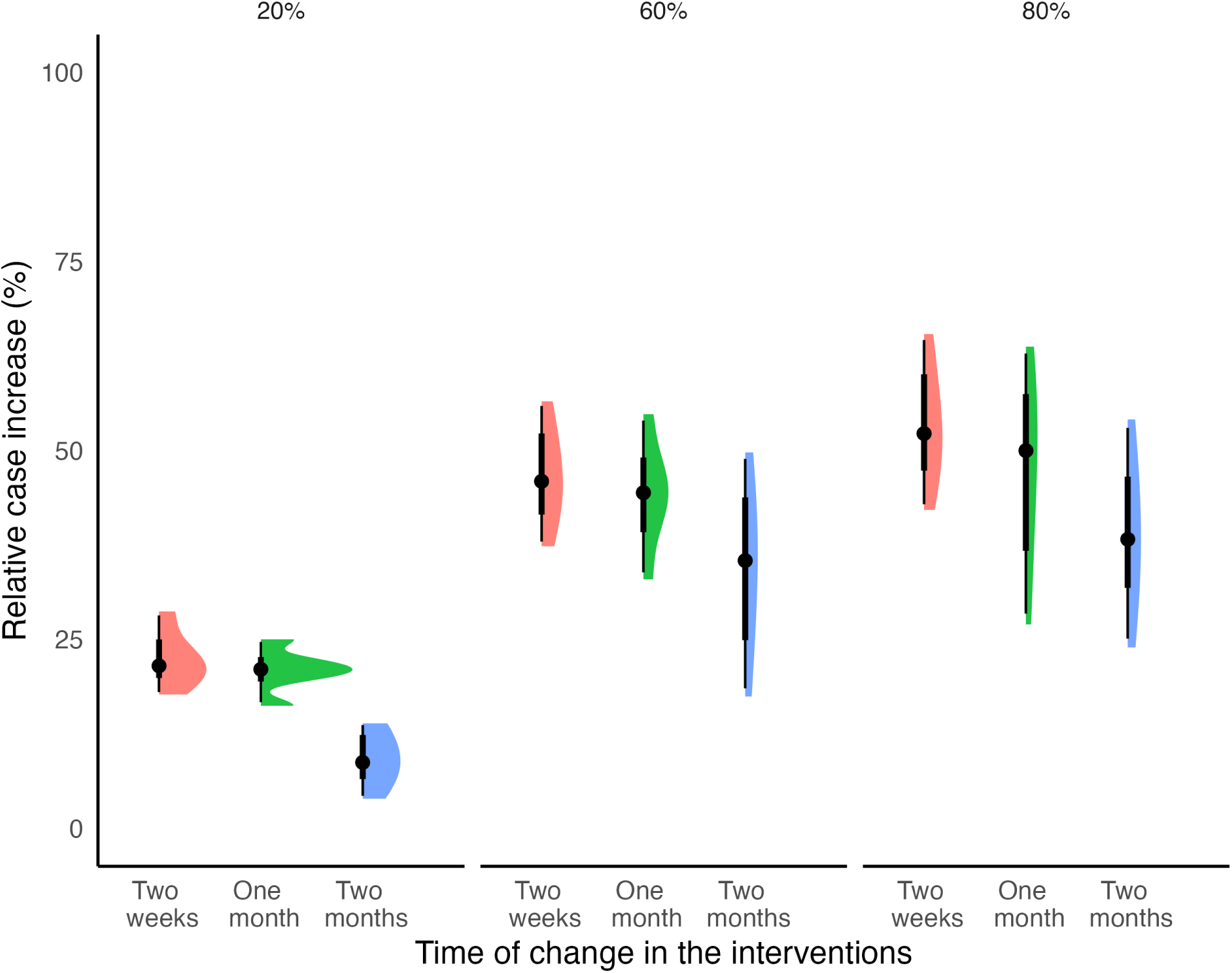
Density interval plots for the percentage increase in the total number of cases summed over all counties for each of the nine counterfactual scenarios in comparison to the observed data. From left to right: an assumed increase in the FoI of 20%, 60%, and 80%. For each assumed increase in the FoI density plots are shown for when interventions are relaxed after two weeks (pink), one month (green), or two months (blue).

## 4 Discussion

In this study, we analysed the dynamics of COVID-19 transmission in rural settings, where population characteristics and occupational patterns stand in stark contrast to the urban environments typically studied in COVID-19 modelling research.

In particular, we studied the transmission dynamics of SARS-CoV-2 within six rural counties of North and South Dakota in the United States. These counties, characterised by small populations of 2500 and low population density, primarily rely on outdoor occupations such as farming, fishing, and forestry (DataUSA, 2022). We estimated that for this group of counties, the proportion of individuals who were tested, notified, and isolated was approximately 0.6 (hyper-parameter for *τ* with uncertainty bounds (from the HPD) from 0.105 to 0.998). We estimated that the reproduction number ranged from 1.1 to 1.7 (quartiles) across counties. Buffalo, SD, had a slightly higher reproduction number [median 1.415 and (1.306, 1.527) quartile] than the other counties. Our estimated reproduction numbers— estimated under the presence of interventions as described—are lower than those globally reported for SARS-CoV-2. However, they are in line with estimates of *R*_0_ in rural areas, likely due to lower contact rates and population densities compared to urban areas(Delamater et al., 2019; Sy et al., 2021).

We conducted a counterfactual analysis to explore the impact of increasing the force of infection by 20%, 60%, and 80% after 2 weeks, 1 month and 2 months. Among the various FoI scenarios considered, we found that an increase of 80% two weeks after the implementation of interventions resulted, as expected, in the maximum relative increase in reported cases. On the other hand, we calculated that if the force of infection had increased by only 20% after two months of implementing public health measures, the relative increase in the total reported cases would not have been substantial.

We used a Bayesian hierarchical analysis to estimate some of the parameters in this study (Gelman, Hill, & Yajima, 2012). Previous work by the authors has shown that using a hierarchical analysis for disease transmission models improves parameter estimation in comparison to conducting parameter estimation independently for each dataset (here a county) (Alahakoon et al., 2022; Alahakoon, McCaw, & Taylor, 2023; Alahakoon, Taylor, & McCaw, 2023). In Alahakoon, Taylor, and McCaw (2023), we investigated the effectiveness of case and contact isolation (at Quarantine Stations) for shipboard outbreaks during the 1918 influenza pandemic. In that work, the outbreaks were modelled as compartmental stochastic processes (continuous time Markov chains). More broadly, in fields such as pharmacometrics— in which ordinary differential equation models are used to study drug and pathogen kinetics—Bayesian hierarchical analysis has a long history dating back to (at least) the early 1990s (Baron, Gillespie, Margossian, et al., 2022; Duffull, Friberg, & Dansirikul, 2007; Smith & Wakefield, 1994). Surprisingly, while Bayesian analyses are common in infectious disease epidemiology, and their use is essentially routine in pharmacometrics, there remains a relative scarcity of studies that apply hierarchical methods to study infectious disease systems described by compartmental models at the epidemiological scale.

Specifically, this study assumed that the transmission rate (*β*) and the proportion of symptomatic individuals who were tested, notified, and isolated immediately (*τ*) followed a hierarchical model and all the other parameters were county-specific. However, we note that this analysis could have been improved if we had conducted a hierarchical analysis that allowed for 1) some parameters to be sampled from a common distribution; 2) some parameters to be county-specific; and 3) other parameters to be the same across all the counties.

We studied only three counties each from South and North Dakota. It may be of interest to study more counties from each state and then conduct a hierarchical analysis with four levels where the additional level is taken for information sharing within the states. Furthermore, it would be interesting to see if similar transmission dynamics are observed in other rural settings.

Another possible future direction for research would be to analyse the transmission dynamics in adjoining counties, in which case the dynamics may be coupled due to the movement of people between counties. In such a setting a hierarchical analysis with a meta-population (or patch) model structure may be of interest.

Additionally, it would be advantageous to extend the transmission model to study the dynamics of the *Omicron* variant where re-infection was observed, the majority of the population was vaccinated, and the non-pharmaceutical intervention measures were largely no longer in place.

## Data Availability

All the codes, data and relevant details are available at https://github.com/PunyaAlahakoon/COVID_19_in_US_rural_counties.git

## Acknowledgements

Punya Alahakoon was supported by a PhD Research Scholarship from the University of Melbourne. P.G. Taylor would like to acknowledge the support of the Australian Research Council via the Centre of Excellence for Mathematical and Statistical Frontiers (ACEMS).

Unless otherwise mentioned, computations were carried out in MATLAB or R across 32 clusters (32 virtual computers). All the computations were carried out by the use of the Nectar Research Cloud (project Infectious Diseases), a collaborative Australian research platform supported by the National Collaborative Research Infrastructure Strategy (NCRIS). All the plots were generated with ggplot2 Wickham (2016) in R. The codes and necessary data are publicly available (see Supplementary Material).

