## supplementary_material for "Stochastic modelling of early-stage COVID-19 epidemic dynamics in rural communities in the United States"

### S1 Data

We downloaded data from `https://github.com/covid19datahub` for each county as CSV files. Each county file consisted of a table where each row represented an observation. The columns included geographical and date identifiers, epidemiological variables (such as cumulative number of confirmed cases, deaths, recovered cases from hospitals, tests, vaccine intake, hospitalisations, people on ventilation, and total population), policy measures (including school closing, workplace closing, cancellation of events, restrictions on gatherings, transport closing, stay at home restrictions, internal and international restrictions, information campaigns, testing policy, contact tracing, facial coverings, vaccination policy, and elderly people protection), and other variables. A graphical representation of some of these variables is included in the GitHub folder (see below for the link).

### S2 Model description

We refer to the model presented in Figure 3 of the main manuscript.

**State description:**

1. $S$ **:** Completely susceptible state.
2. $V_1$ **:** The first dose of the vaccine has been received.
3. $V_2$ **:** The second dose of the vaccine has been received.
4. $E$ **:** Exposed state.
5. $A_1$ **:** First asymptomatic state.
6. $A_2$ **:** Second asymptomatic state.
7. $R_A$ **:** Recovery of an asymptomatic person.
8. $P$ **:** Pre-symptomatic state.
9. $I$ **:** First symptomatic infectious state.
10. $C$**:** Symptomatic infectious and positive tested for COVID-19. It is assumed that individuals in this state will immediately isolate themselves once tested positive).
11. $I_2$ **:** Second symptomatic infectious state. It is assumed that individuals in this state do not get tested for COVID-19 and therefore will not be recorded as positive cases.

*

12. $R$ **:** Recovery of symptomatically infectious person.

**Parameter description:**

1. $\lambda$ : Force of infection where,

$$\lambda = \beta\Big[\frac{A_1 + A_2 + P + I + I_2}{N-1}\Big], \tag{S.1}$$

   and $N$ is the population size of the county and $\beta$ is the transmission rate.
2. $\nu_1$: The rate at which fully susceptible people receive the first dose of the COVID-19 vaccine.
3. $\nu_2$: The rate at which people received the first dose receive their second dose.
4. $\lambda(1-\zeta_1)$: Rate at which individuals who are partially vaccinated (dose 1) become infectious and $\zeta_1 \leq 1$.
5. $\lambda(1-\zeta_2)$: Rate at which individuals who are fully vaccinated (dose 2) become infectious and $\zeta_2 \leq 1$.
6. $\delta(1-\omega)$: The rate at which an exposed person first becomes asymptomatic and $\omega \leq 1$.
7. $\epsilon_1$: The rate at which a person in the first asymptomatic state moves to the second asymptomatic state.
8. $\epsilon_2$: The rate at which a person in the second asymptomatic state recovers.
9. $\delta\omega$: The rate at which an exposed person becomes pre-symptomatic.
10. $\alpha$: The rate at which a pre-symptomatic person first becomes symptomatically infectious.
11. $\tau\gamma_1$: Rate at which a person in the first infectious state tests positive for COVID-19 and $\tau \leq 1$.
12. $(1-\tau)\gamma_1$: Rate at which a person in the first infectious state moves to the second infectious state without getting tested and $\tau \leq 1$.
13. $\gamma_2$ : The rate at which a person in either $C$ or $I_2$ states recovers.
14. $d$ : Death rate of COVID1-19 in the county.
15. $b$ : Rate at which COVID-19 is re-introduced to the county.

**Model assumptions:**

1. Individuals in state $C$ will not contribute to the transmission of the disease as they remain in isolation once they are tested positive.
2. The asymptomatic and symptomatic transmissions are the same.
3. The transmission rate will be constant across the time period.
4. The vaccine protection is achieved after 14 days of the first and second doses.

### S3 Parameter estimation

We used the two-step algorithm of Alahakoon, McCaw, and Taylor (2022) to estimate parameters under a hierarchical framework. Step 1(a) of the algorithm conducts parameter estimation by considering each outbreak independently. We used the ABC-SMC algorithm of Toni, Welch, Strelkowa, Ipsen, and Stumpf (2009). Details are explained below.

Then we estimated the hyper-parameters under a hierarchical framework. We assumed that $\beta_k$ and $\tau_k$ $(k = 1, 2, \ldots, 6)$ are sampled from a truncated multivariate normal distribution (that is, the conditional prior distribution).

The prior distributions and the conditional prior distributions are:

$$
\begin{aligned}
\Psi_\beta &\sim \text{Uniform}(0.00001, 2)\\
\sigma_\beta &\sim \text{Uniform}(0, 2)\\
\Psi_\tau &\sim \text{Uniform}(0.00001, 1)\\
\sigma_\tau &\sim \text{Uniform}(0, 2)\\
R &\sim \text{LKJcorr}(2) \quad \text{(Prior for correlation matrix)}\\
(\beta_k, \tau_k) &\sim \text{Truncated Multivariate Normal}(\boldsymbol{\Psi}, \Sigma, \mathbf{a}, \mathbf{b}),
\end{aligned}
$$

where,

$$
R = \begin{bmatrix} 1 & \rho_{12} \\ \rho_{21} & 1 \end{bmatrix} \tag{S.2}
$$

the covariance matrix

$$
\Sigma = \begin{bmatrix} \sigma_\beta & 0 \\ 0 & \sigma_\tau \end{bmatrix} R \begin{bmatrix} \sigma_\beta & 0 \\ 0 & \sigma_\tau \end{bmatrix} \tag{S.3}
$$

and $\mathbf{a} = (0.00001, 0.00001)$ and $\mathbf{b} = (2, 1)$. The relevant R codes are included in GitHub.

Once hyperparameters were estimated, the parameters of the counties using an ABC algorithm. We proposed the transmission rates using the conditional prior, Truncated Multivariate Normal($\boldsymbol{\Psi}, \Sigma, \mathbf{a}, \mathbf{b}$), with the estimated hyper-parameters. All the other parameters were sampled by perturbing from the last generation of the marginal posterior distribution from Step 1(a). In this step, we used the final generations' tolerance values of the ABC-SMC algorithm we used in the first step.

#### S3.1 parameter estimation by considering each outbreak independently

ABC-SMC algorithm was used to estimate the parameters. We used the prior distributions presented in Table S1 for all the counties.

Table S1: Prior distributions

| Parameter | Prior distribution |
|---|---|
| $\beta$ | Uniform (0.00001,2) |
| $\lambda(1-\zeta_1), \lambda(1-\zeta_2)$ | Uniform (0.0001,1) |
| $\nu_1$ | Uniform (0.0000001, 0.05) |
| $\nu_2$ | Uniform (0.0000001, 0.1) |
| $1/\delta(1-\omega)$ | lognormal (0.4039,0.6) |
| $1/\epsilon_1$ | lognormal (0.6619,0.25) |
| $1/\epsilon_2$ | lognormal (-0.3, 0.5) |
| $1/\delta\omega$ | lognormal (1.125,0.35) |
| $1/\alpha$ | lognormal (0.635,0.35) |
| $\tau$ | Uniform (0.00001,1) |
| $1/\tau\gamma_1$ | lognormal (1.485,0.5) |
| $1/\gamma_2$ | lognormal (1.485,0.5) |
| $b$ | Uniform (0, 1) |
| $d$ | Beta (1.5, 30) |

The ABC-SMC algorithm used the following distance criteria across three or four generations (depending on the county).

1. Euclidean distance of the observed and generated daily new cases.
2. Absolute difference of the cumulative sums of deaths at the end of the considered time period between the observed and generated epidemic curves.
3. The absolute difference between the generated and observed cumulative sums of dose 1 vaccinations. Time series data from the day the vaccine became available to the first non-zero count are excluded.
4. The absolute difference between the generated and observed cumulative sums of dose 2 vaccinations. Time series data from the day the vaccine became available to the first non-zero count are excluded.
5. The absolute difference between the cumulative sums of the generated and observed data for the initial phase of the epidemic (i.e., before the major outbreak is assumed to have started).
6. The absolute difference between the cumulative sums of the generated and observed data for the final phase of the epidemic (i.e., after the major outbreak is assumed to have subsided).
7. The absolute difference between the cumulative sum of the generated dose 1 vaccinations from the date of vaccine availability to the date of data availability and the dose 1 vaccine uptake recorded on the first date (considered due to the unusual peak in the vaccine data).
8. The absolute difference between the cumulative sum of the generated dose 2 vaccinations from the date of vaccine availability to the date of data availability and the dose 1 vaccine uptake recorded on the first date (considered due to the unusual peak in the vaccine data).

The tables below represent tolerance values used for the six counties.

Table S2: Tolerance values for Miner, SD

| **Generation** | **Distance criteria** | | | | | | | |
|---|---|---|---|---|---|---|---|---|
| | **1** | **2** | **3** | **4** | **5** | **6** | **7** | **8** |
| 1 | 37 | 8 | 912 | 881 | 37 | 15 | 246 | 74 |
| 2 | 35 | 6 | 800 | 700 | 30 | 13 | 200 | 70 |
| 3 | 33 | 5 | 700 | 600 | 20 | 11 | 150 | 60 |

Table S3: Tolerance values for Buffalo, SD

| **Generation** | **Distance criteria** | | | | | | | |
|---|---|---|---|---|---|---|---|---|
| | **1** | **2** | **3** | **4** | **5** | **6** | **7** | **8** |
| 1 | 37 | 8 | 912 | 881 | 37 | 15 | 246 | 74 |
| 2 | 35 | 6 | 800 | 700 | 30 | 13 | 200 | 70 |
| 3 | 33 | 5 | 700 | 600 | 20 | 11 | 150 | 60 |

Table S4: Tolerance values for Faulk, SD

| **Generation** | **Distance criteria** | | | | | | | |
|---|---|---|---|---|---|---|---|---|
| | **1** | **2** | **3** | **4** | **5** | **6** | **7** | **8** |
| 1 | 47 | 12 | 752 | 876 | 11 | 50 | 195 | 82 |
| 2 | 45 | 10 | 600 | 700 | 10 | 45 | 170 | 70 |
| 3 | 43 | 8 | 500 | 600 | 9 | 40 | 150 | 60 |

Table S5: Tolerance values for Towner, ND

| Generation | Distance criteria | | | | | | | |
|---|---|---|---|---|---|---|---|---|
| | 1 | 2 | 3 | 4 | 5 | 6 | 7 | 8 |
| 1 | 38 | 10 | 820 | 787 | 27 | 18 | 229 | 61 |
| 2 | 36 | 8 | 750 | 700 | 20 | 15 | 150 | 50 |
| 3 | 34 | 6 | 650 | 650 | 15 | 12 | 130 | 45 |
| 4 | 32 | 5 | 550 | 600 | 10 | 8 | 120 | 40 |

Table S6: Tolerance values for Eddy, ND

| Generation | Distance criteria | | | | | | | |
|---|---|---|---|---|---|---|---|---|
| | 1 | 2 | 3 | 4 | 5 | 6 | 7 | 8 |
| 1 | 64 | 5 | 973 | 916 | 17 | 52 | 302 | 26 |
| 2 | 63 | 4 | 850 | 850 | 15 | 45 | 200 | 20 |
| 3 | 60 | 3 | 800 | 800 | 10 | 35 | 170 | 17 |

Table S7: Tolerance values for Golden Valley, ND

| Generation | Distance criteria | | | | | | | |
|---|---|---|---|---|---|---|---|---|
| | 1 | 2 | 3 | 4 | 5 | 6 | 7 | 8 |
| 1 | 33 | 2 | 445 | 362 | 20 | 37 | 147 | 36 |
| 2 | 32 | 2 | 350 | 300 | 15 | 35 | 140 | 30 |
| 3 | 31 | 2 | 300 | 250 | 10 | 34 | 100 | 25 |
| 4 | 30 | 2 | 250 | 200 | 8 | 33 | 70 | 20 |

### S4 Hyper parameters

#### Hyper mean

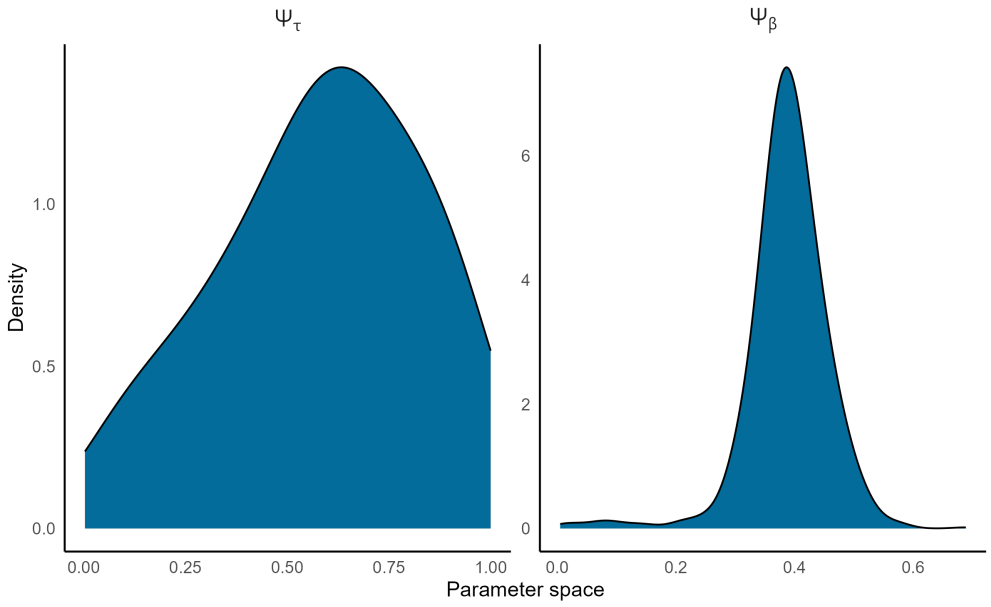


Figure S1: Marginal posterior distributions of the hyper-parameters $\psi_\beta$ and $\psi_\tau$

Table S8: Posterior medians and 95% HPD intervals of the hyper-parameters $\psi_\beta$ and $\psi_\tau$

| Parameter | Posterior median | 95% HPD interval |
|---|---|---|
| $\psi_\beta$ | 0.391 | (0.285, 0.524) |
| $\psi_\tau$ | 0.606 | (0.105, 0.992) |

**Standard deviation**

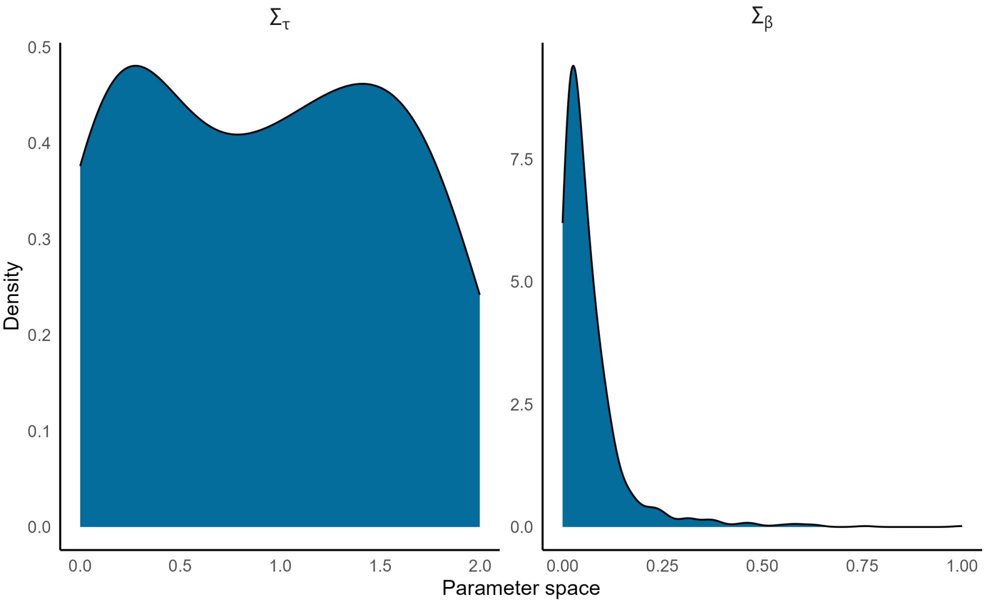


Figure S2: Marginal posterior distributions of the hyper-parameters $\sigma_\beta$ and $\sigma_\tau$

Table S9: Posterior medians and 95% HPD intervals of the hyper-parameters $\sigma_\beta$ and $\sigma_\tau$

| Parameter | Posterior median | 95% HPD interval |
|---|---|---|
| $\sigma_\beta$ | 0.416 | (0, 0.227) |
| $\sigma_\tau$ | 0.930 | (0.001, 1.886) |

**County-specific $\tau$**

Table S10: Posterior medians and 95% HPD intervals for $\tau$, the proportion of symptomatic individuals who were tested, notified, and isolated immediately

| County | $\tau$ Posterior median (95% HPD interval) |
|---|---|
| Miner, SD | 0.604 (0.277, 0.999) |
| Buffalo, SD | 0.626 (0.281, 0.992) |
| Faulk, SD | 0.590 (0.217, 0.971) |
| Towner, ND | 0.585 (0.260, 0.968) |
| Eddy, ND | 0.637 (0.320, 0.999) |
| Golden Valley, ND | 0.617 (0.286, 0.998) |

**Estimated county-specific $R_0$**

Table S11: Estimated $R_0$s of the counties

| County | Median $R_0$ (25, 75)% quantile interval |
|---|---|
| Miner, SD | 1.204 (1.100, 1.305) |
| Buffalo, SD | 1.415 (1.306, 1.527) |
| Faulk, SD | 1.330 (1.243, 1.305) |
| Towner, ND | 1.320 (1.230, 1.408) |
| Eddy, ND | 1.379 (1.202, 1.627) |
| Golden Valley, ND | 1.192 (1.095, 1.300) |

#### S4.1 Comparison of parameters with and without a hierarchical analysis

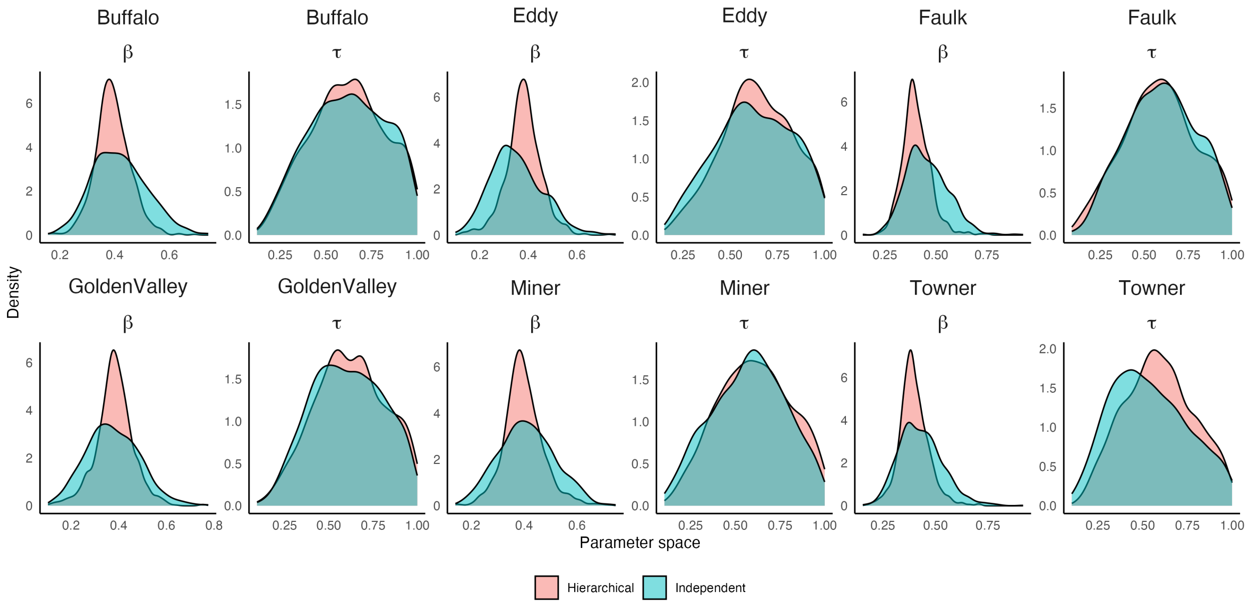


Figure S3: Marginal posterior distributions of the hyper-parameters $\sigma_\beta$ and $\sigma_\tau$

### S5 Re-sampled paths and re-sampled paths of vaccine intake

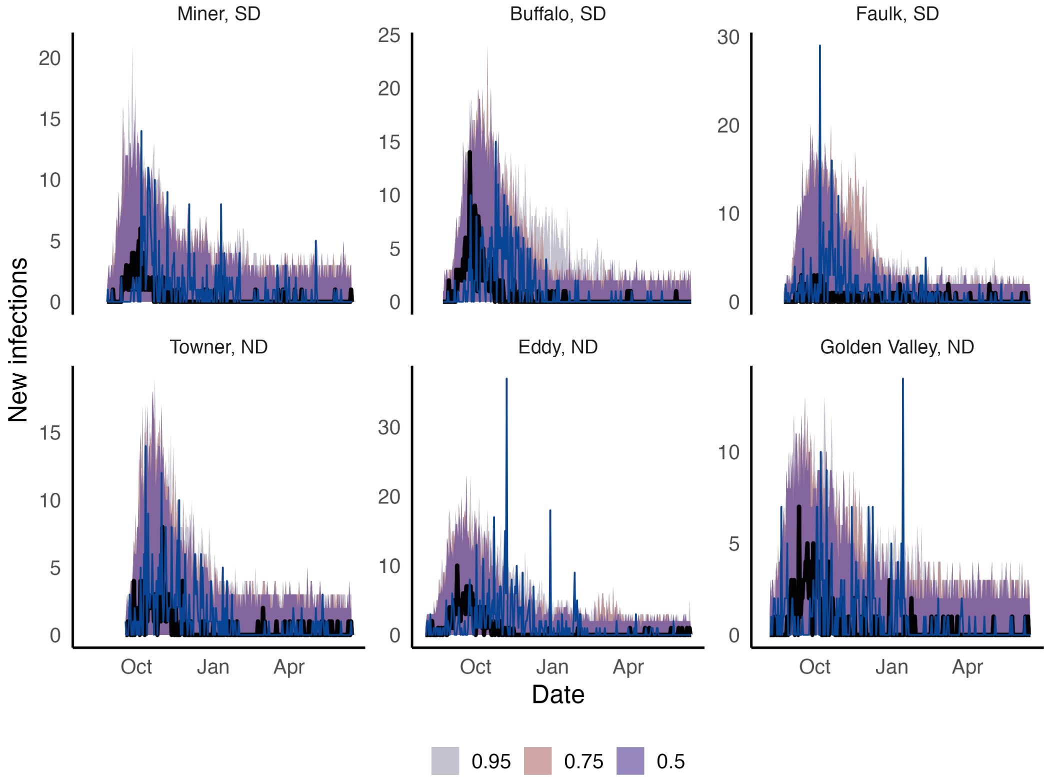


Figure S4: Re-sampled for the new infections. The black line is the median predicted path, the 50%, 75%, and 95% curve-wise intervals are presented in purple, pink and grey ribbons, respectively. The blue lines show new infections.

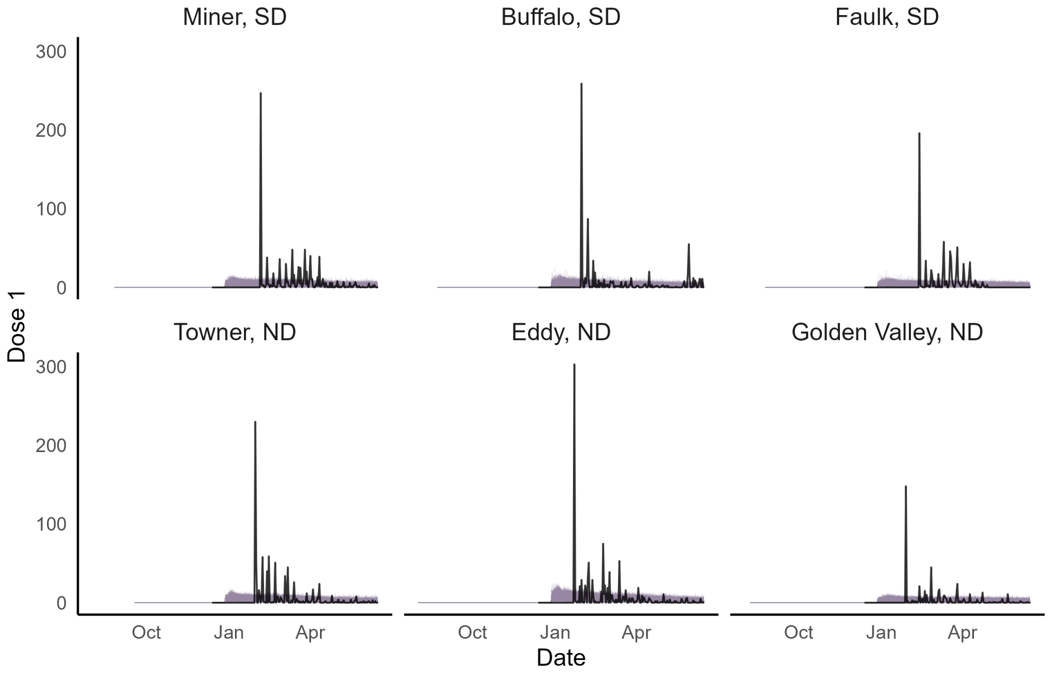


Figure S5: Re-sampled paths for Dose 1

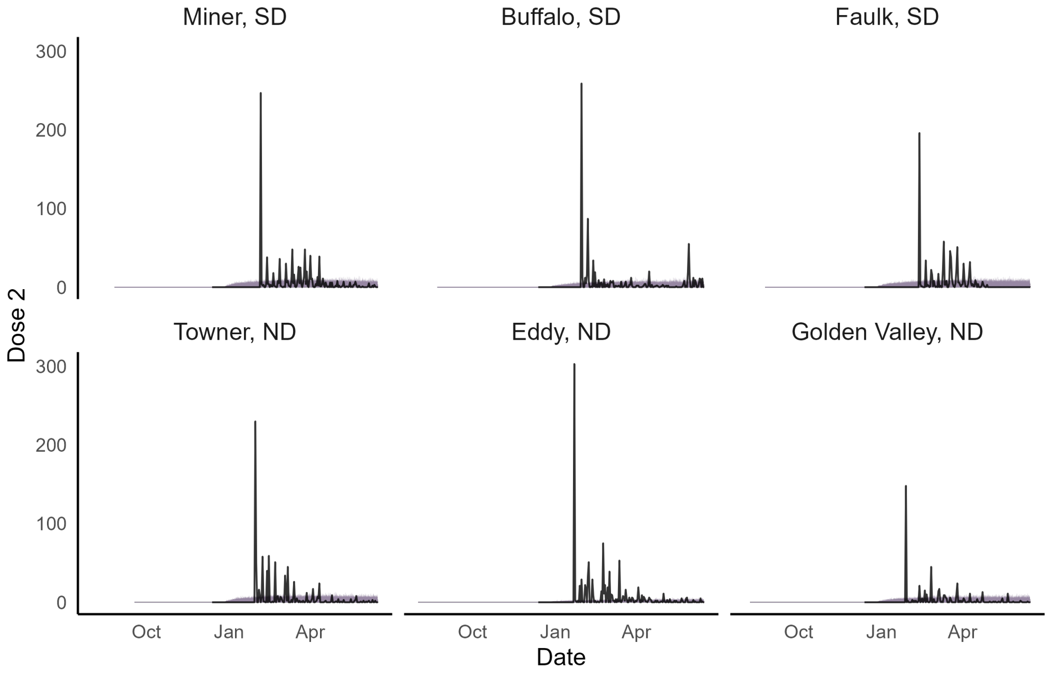


Figure S6: Re-sampled paths for Dose 2

### S6 All posterior distributions under a hierarchical framework

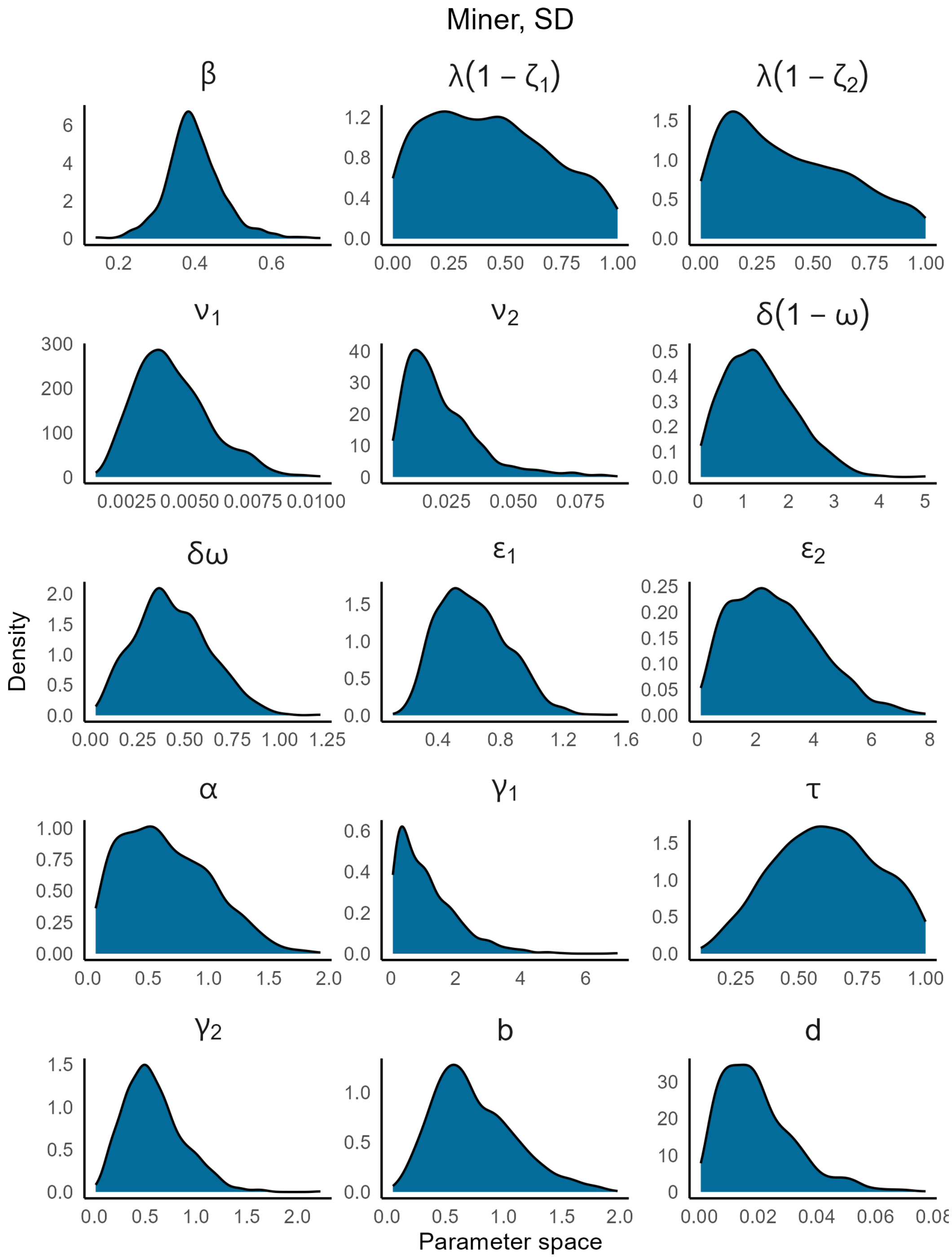


Figure S7: Marginal posterior distributions of Miner, SD

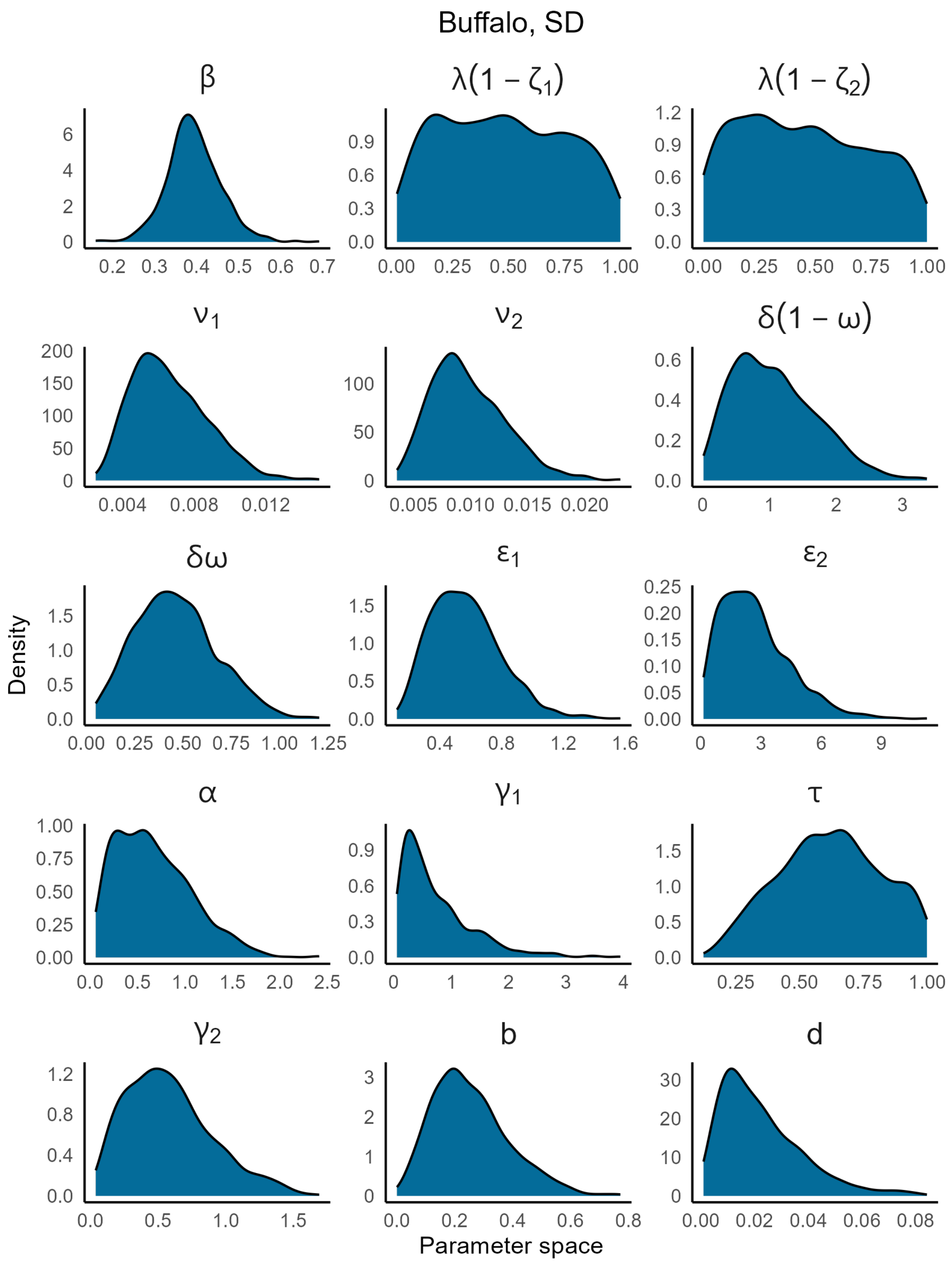


Figure S8: Marginal posterior distributions of Buffalo, SD

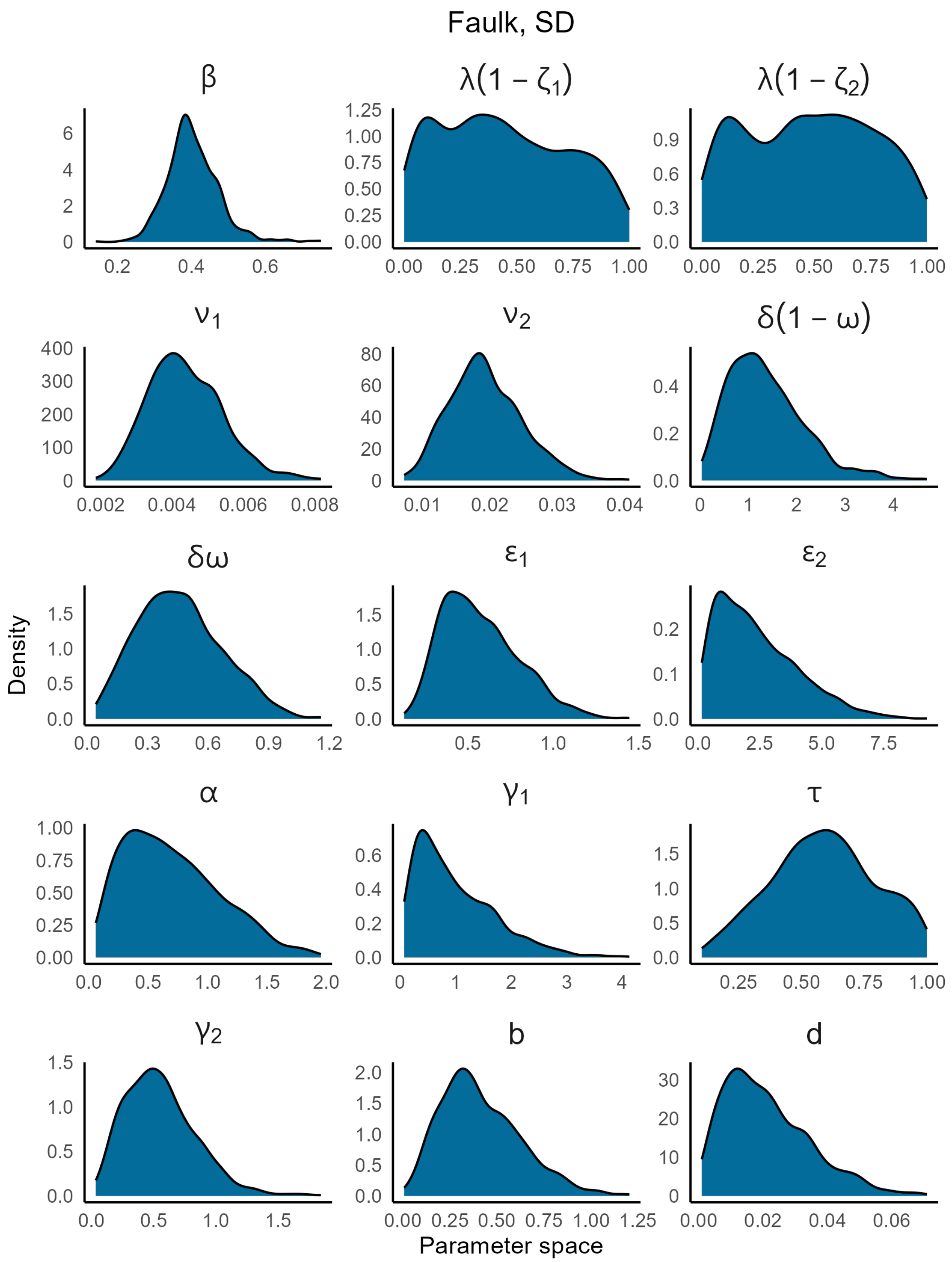


Figure S9: Marginal posterior distributions of Faulk, SD

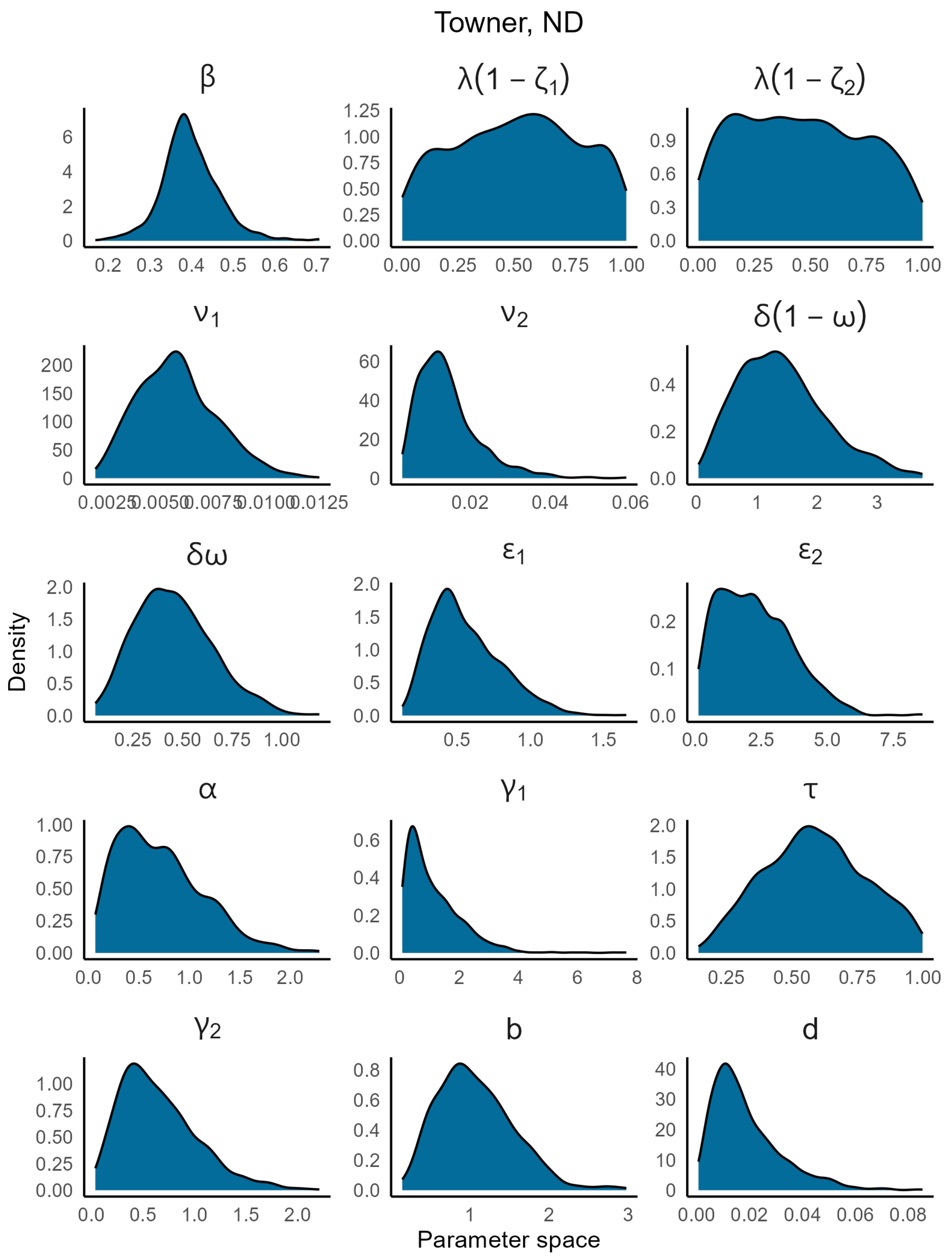


Figure S10: Marginal posterior distributions of Towner, ND

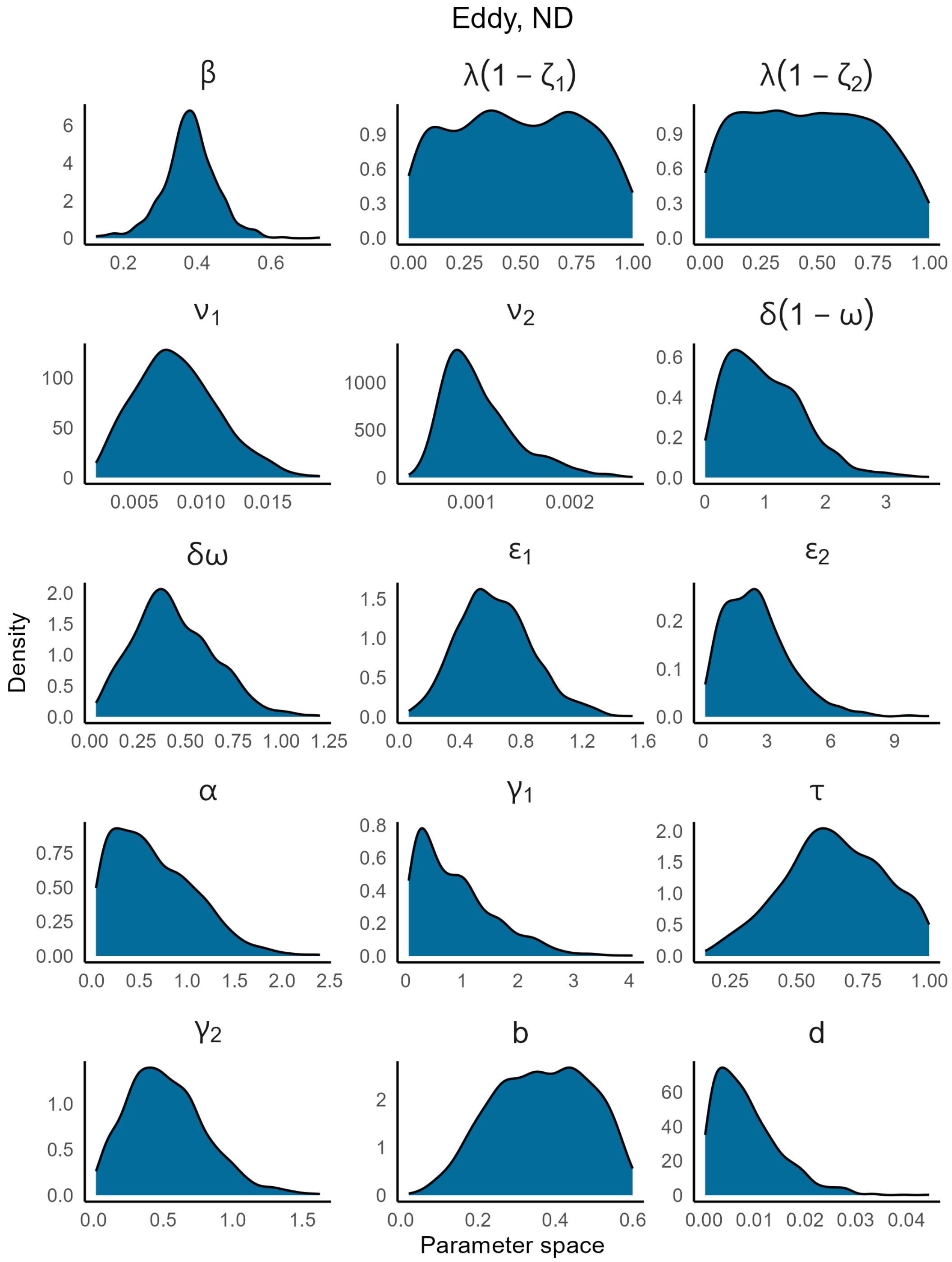


Figure S11: Marginal posterior distributions of Eddy, ND

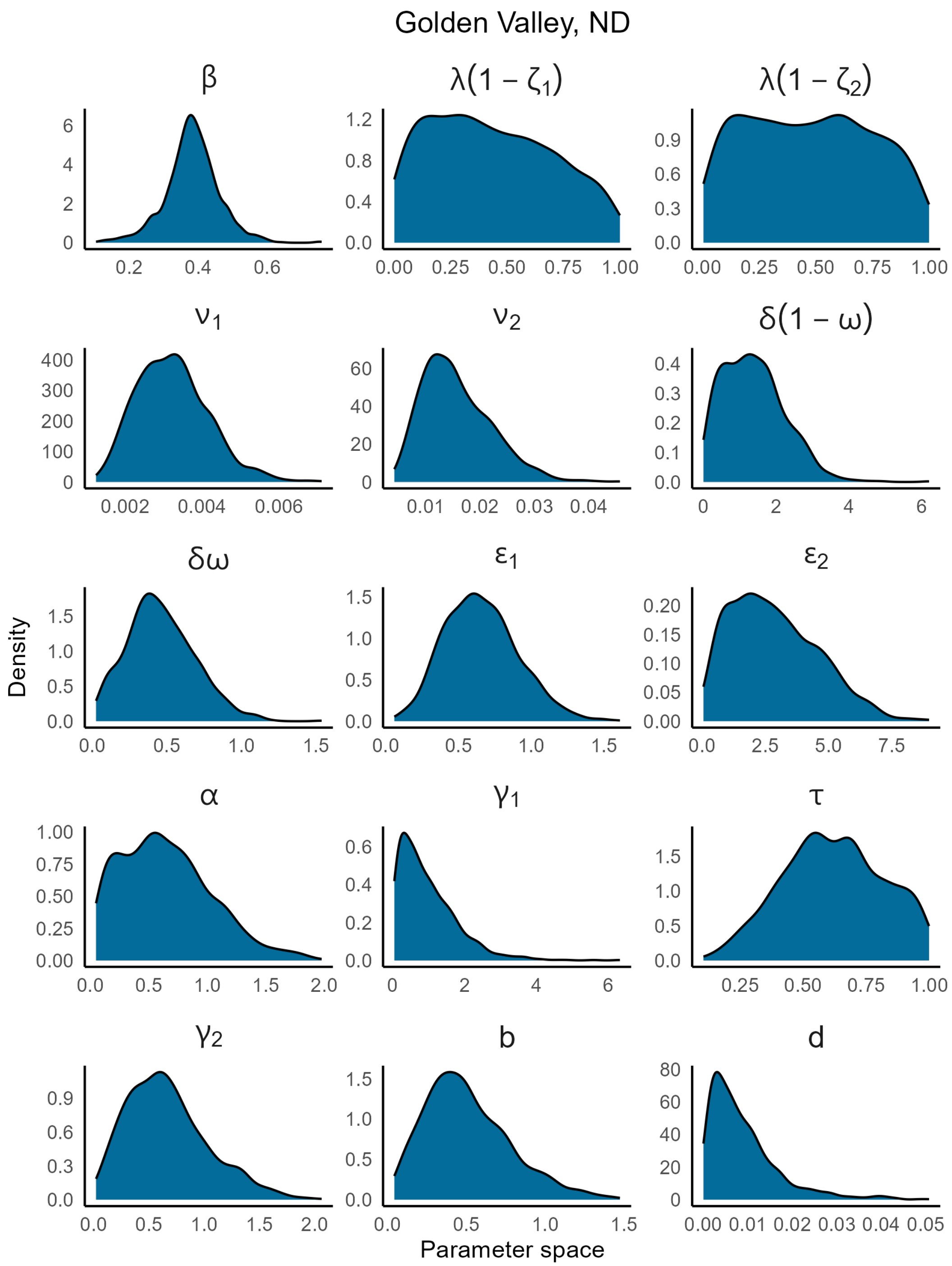


Figure S12: Marginal posterior distributions of Golden Valley, ND

### S7 Mobility data analysis

CDC link used: `https://covid.cdc.gov/covid-data-tracker/#mobility` Google mobility data analysis `https://www.google.com/covid19/mobility/` can be found in the GitHub folder (link provided below).

### S8 Counterfactuals

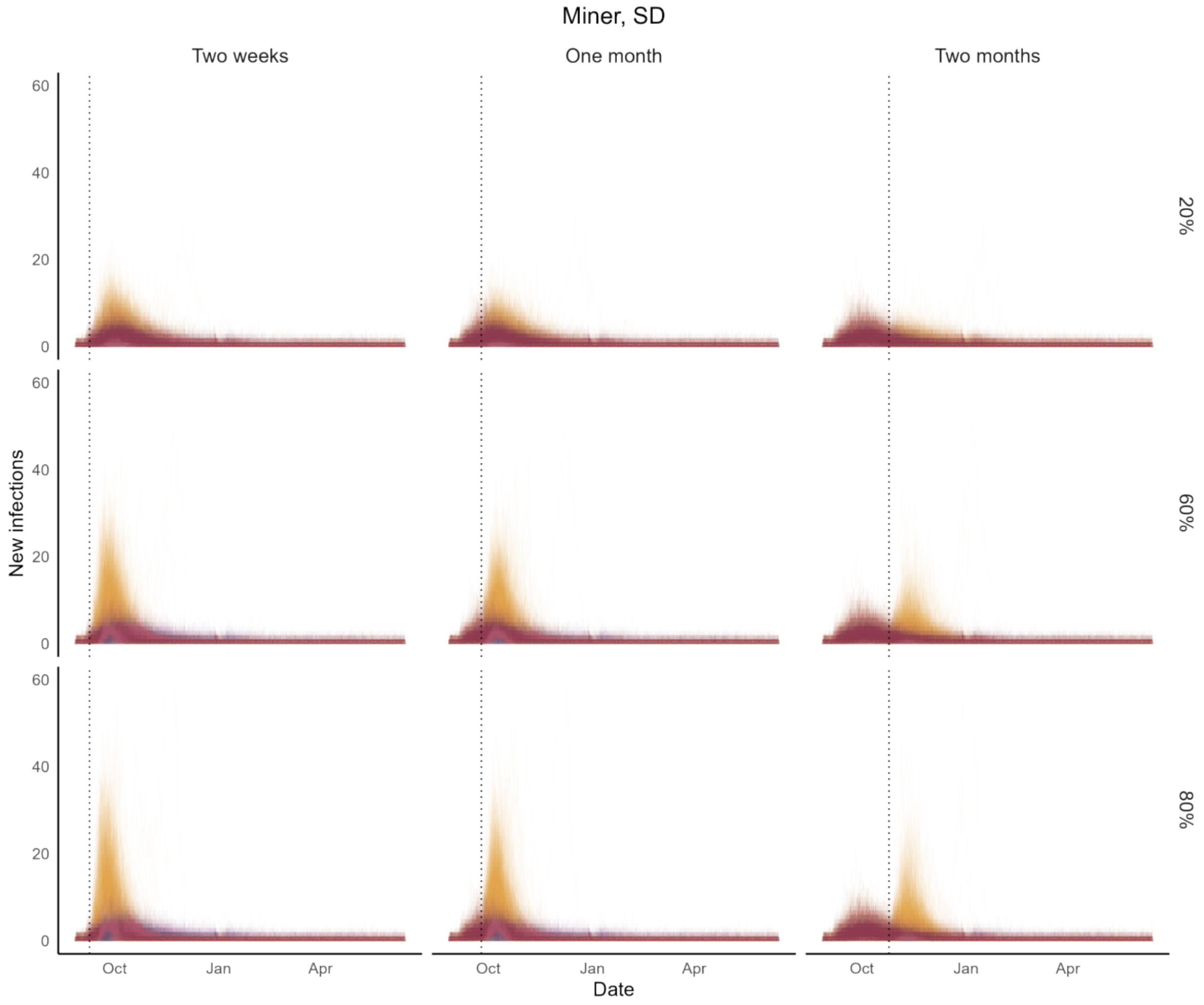


Figure S13: Impact of COVID-19 on Miner, SD had the FoI increased by 20% (first row), 60% (second row), and 80% (third row)after 2 weeks (first column), one month (second column), and two months (third column). Purple paths are predicted paths with estimated FoI. Yellow sample paths are counterfactuals

Table S12: Increase of total cases if FoI was increased at two weeks, one month, and two months for Miner, SD

| **Increase of FOI** | **Time of change in the interventions** | **Median** | **25% quantile** | **75% quantile** | **Relative case increase %** | **Relative case increase 25% quantile** | **Relative case increase 75% quantile** |
|---|---|---|---|---|---|---|---|
| 20% | Two weeks | 42.5 | 26 | 65 | 24.33 | 14.78 | 38.20 |
| 20% | One month | 39 | 24 | 59 | 22.26 | 14.06 | 34.20 |
| 20% | Two months | 16 | 4 | 35 | 9.88 | 2.25 | 20.70 |
| 60% | Two weeks | 91 | 64 | 131 | 51.47 | 36.69 | 76.72 |
| 60% | One month | 84.5 | 58 | 125 | 48.03 | 33.67 | 73.51 |
| 60% | Two months | 60 | 36 | 103 | 36.01 | 20.34 | 60.47 |
| 80% | Two weeks | 105 | 74 | 149.25 | 59.13 | 43.03 | 88.14 |
| 80% | One month | 99 | 69 | 146.25 | 56.34 | 39.57 | 84.73 |
| 80% | Two months | 80 | 48 | 126.25 | 45.20 | 26.56 | 74.85 |

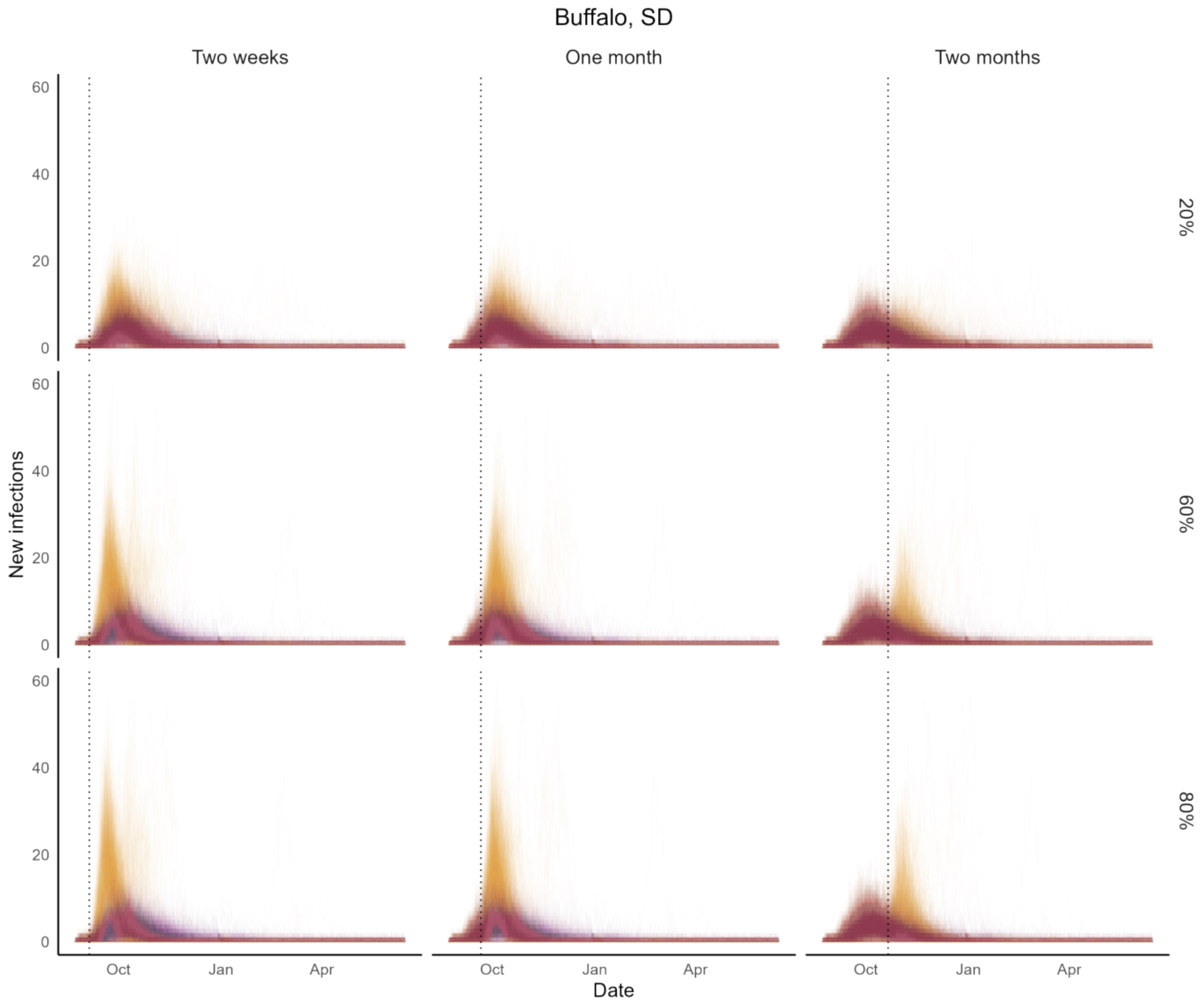


Figure S14: Impact of COVID-19 on Buffalo, SD had the FoI increased by 20% (first row), 60% (second row), and 80% (third row)after 2 weeks (first column), one month (second column), and two months (third column). Purple paths are predicted paths with estimated FoI. Yellow sample paths are counterfactuals

Table S13: Increase of total cases if FoI was increased at two weeks, one month, and two months for Buffalo, SD

| **Increase of FOI** | **Time of change in the interventions** | **Median** | **25% quantile** | **75% quantile** | **Relative case increase %** | **Relative case increase 25% quantile** | **Relative case increase 75% quantile** |
|---|---|---|---|---|---|---|---|
| 20% | Two weeks | 45.5 | 28 | 66 | 20.67 | 12.95 | 29.96 |
| 20% | One month | 44 | 29 | 62 | 20.00 | 13.25 | 29.17 |
| 20% | Two months | 15 | 4 | 36.25 | 6.99 | 1.86 | 16.96 |
| 60% | Two weeks | 93 | 68 | 127 | 42.25 | 31.40 | 57.59 |
| 60% | One month | 90 | 65 | 124 | 40.29 | 29.77 | 56.33 |
| 60% | Two months | 58 | 32 | 93 | 26.21 | 14.60 | 43.62 |
| 80% | Two weeks | 108 | 78 | 145 | 48.24 | 36.85 | 65.46 |
| 80% | One month | 103 | 76 | 142.25 | 47.06 | 35.44 | 63.96 |
| 80% | Two months | 73 | 45 | 114.25 | 33.20 | 20.60 | 53.27 |

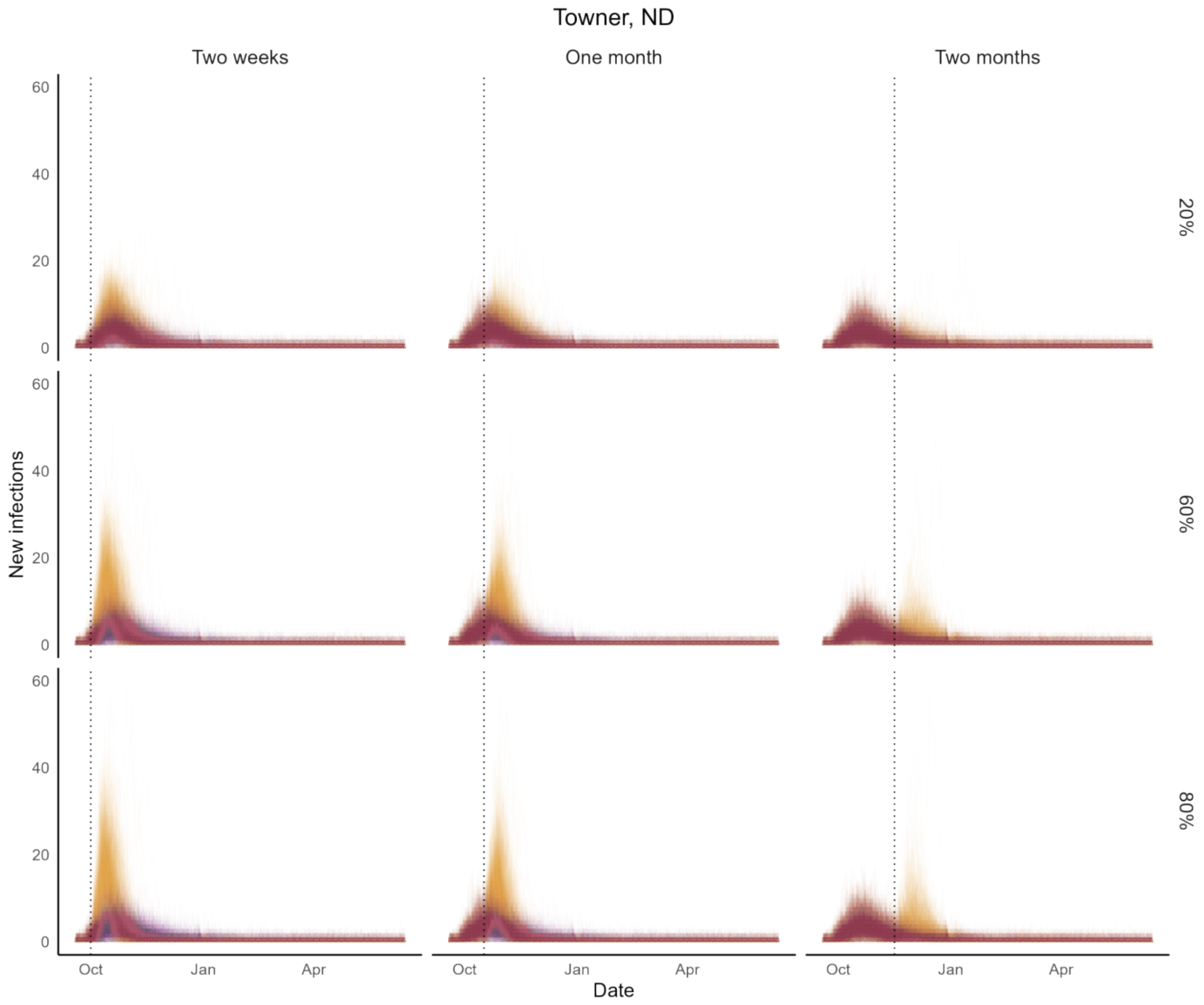


Figure S15: Impact of COVID-19 on Towner, ND had the FoI increased by 20% (first row), 60% (second row), and 80% (third row)after 2 weeks (first column), one month (second column), and two months (third column). Purple paths are predicted paths with estimated FoI. Yellow sample paths are counterfactuals

Table S14: Increase of total cases if FoI was increased at two weeks, one month, and two months for Towner, ND

| **Increase of FOI** | **Time of change in the interventions** | **Median** | **25% quantile** | **75% quantile** | **Relative case increase %** | **Relative case increase 25% quantile** | **Relative case increase 75% quantile** |
|---|---|---|---|---|---|---|---|
| 20% | Two weeks | 37 | 23 | 52 | 17.71 | 11.38 | 25.14 |
| 20% | One month | 33 | 20 | 50 | 16.23 | 9.56 | 23.60 |
| 20% | Two months | 8 | 0 | 18 | 3.93 | 0.00 | 8.90 |
| 60% | Two weeks | 77 | 56 | 104 | 37.32 | 27.74 | 48.70 |
| 60% | One month | 69 | 50 | 98 | 32.95 | 25.19 | 45.87 |
| 60% | Two months | 37 | 23 | 59 | 17.45 | 11.34 | 27.76 |
| 80% | Two weeks | 88 | 64 | 119 | 42.12 | 31.84 | 54.81 |
| 80% | One month | 80 | 58 | 111 | 38.45 | 28.43 | 51.69 |
| 80% | Two months | 50 | 34 | 77 | 23.94 | 16.65 | 36.21 |

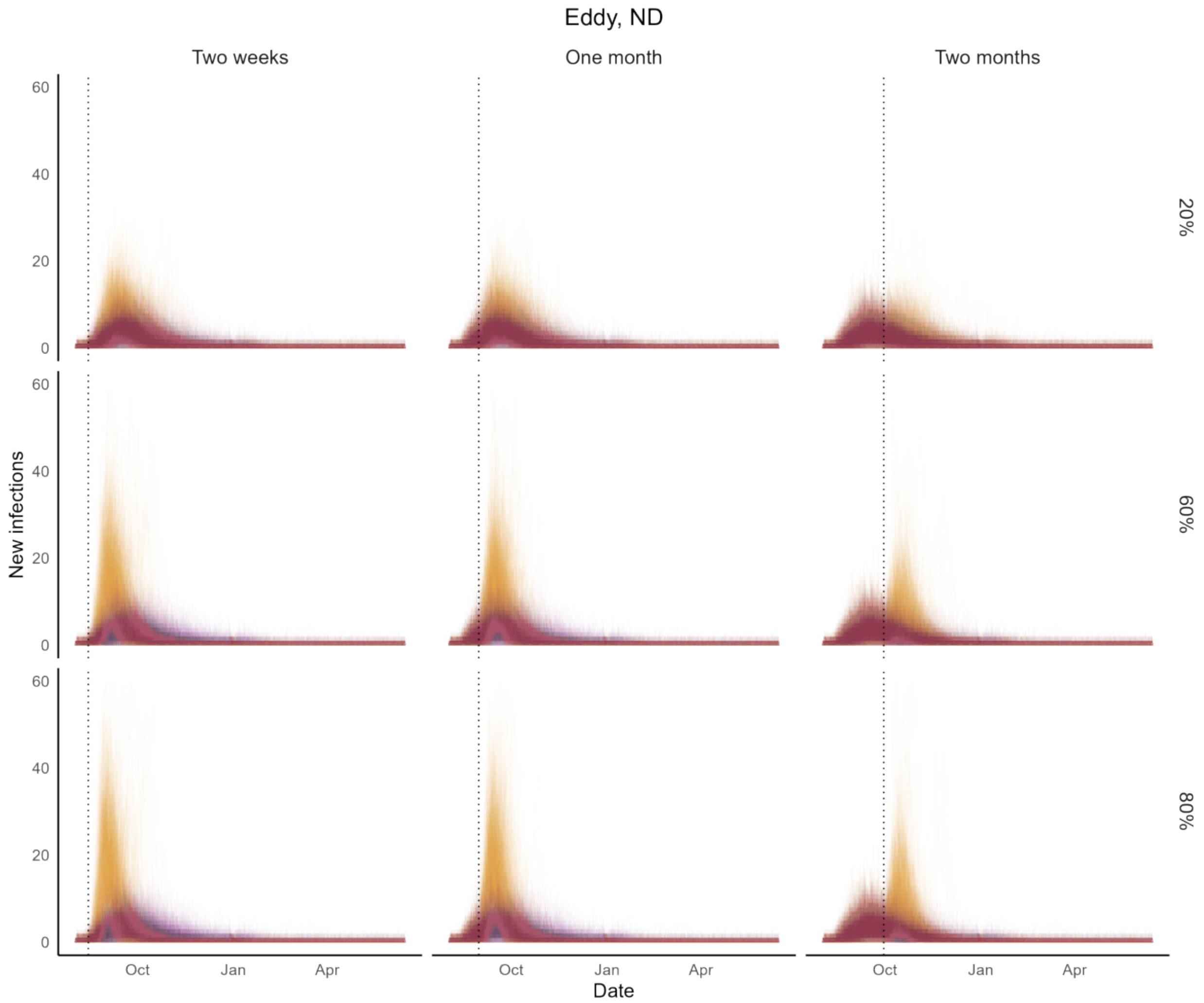


Figure S16: Impact of COVID-19 on Eddy, ND had the FoI increased by 20% (first row), 60% (second row), and 80% (third row)after 2 weeks (first column), one month (second column), and two months (third column). Purple paths are predicted paths with estimated FoI. Yellow sample paths are counterfactuals

Table S15: Increase of total cases if FoI was increased at two weeks, one month, and two months for Eddy, ND

| Increase of FOI | Time of change in the interventions | Median | 25% quantile | 75% quantile | Relative case increase % | Relative case increase 25% quantile | Relative case increase 75% quantile |
|---|---|---|---|---|---|---|---|
| 20% | Two weeks | 58 | 38 | 84 | 22.34 | 14.60 | 32.00 |
| 20% | One month | 56 | 37 | 80 | 21.43 | 14.56 | 31.47 |
| 20% | Two months | 30.5 | 13.75 | 55 | 12.10 | 5.29 | 21.35 |
| 60% | Two weeks | 123.5 | 86.75 | 172 | 46.87 | 35.07 | 65.91 |
| 60% | One month | 120 | 84 | 167.25 | 45.66 | 34.12 | 63.39 |
| 60% | Two months | 90.5 | 56 | 141.25 | 34.84 | 22.66 | 52.75 |
| 80% | Two weeks | 138 | 100 | 197.25 | 53.20 | 39.53 | 74.35 |
| 80% | One month | 136.5 | 100 | 193.25 | 52.88 | 40.03 | 73.72 |
| 80% | Two months | 111 | 72 | 168.25 | 42.14 | 29.57 | 63.57 |

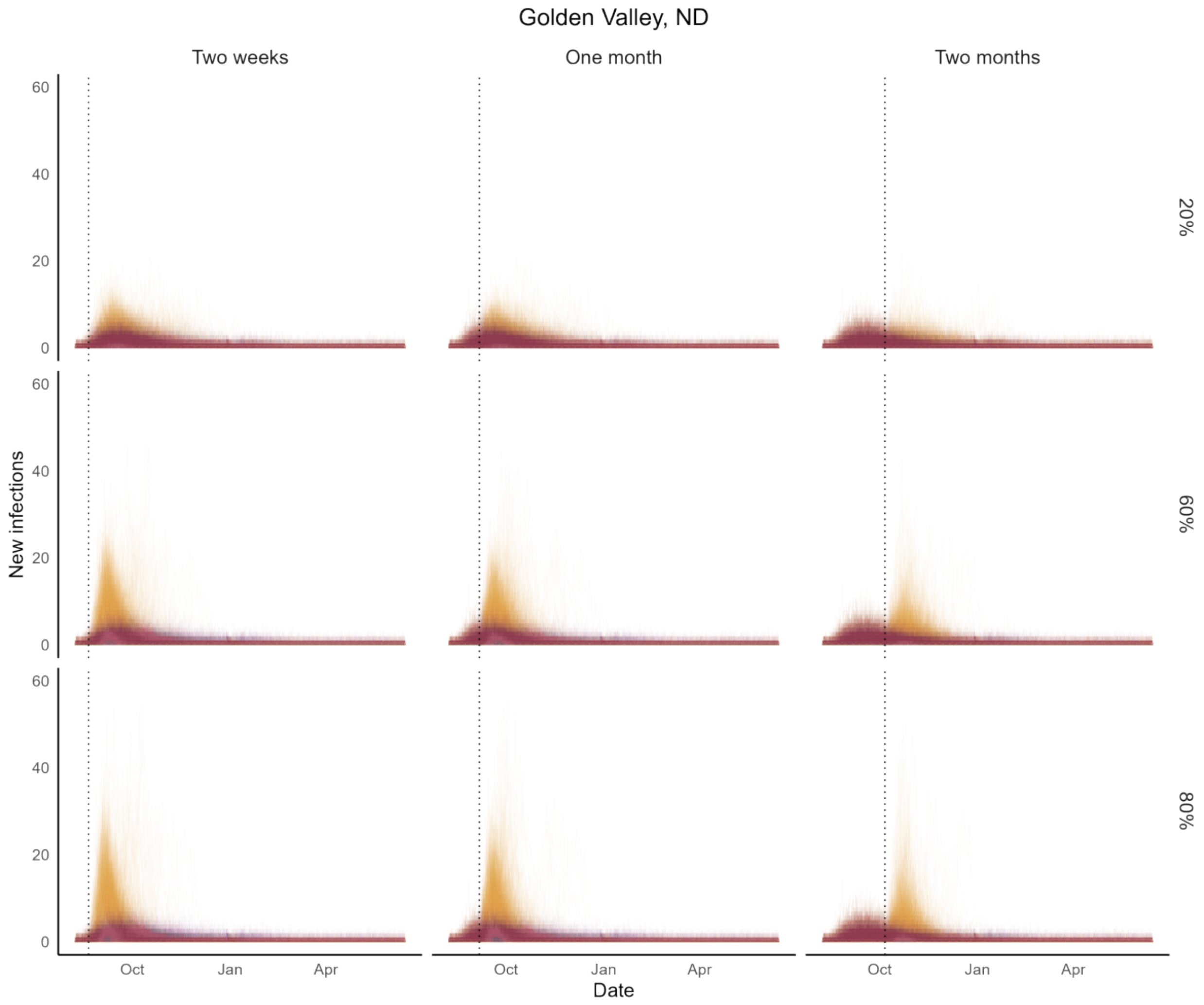


Figure S17: Impact of COVID-19 on Golden Valley, ND had the FoI increased by 20% (first row), 60% (second row), and 80% (third row)after 2 weeks (first column), one month (second column), and two months (third column). Purple paths are predicted paths with estimated FoI. Yellow sample paths are counterfactuals

Table S16: Increase of total cases if FoI was increased at two weeks, one month, and two months for

| **Increase of FOI** | **Time of change in the interventions** | **Median** | **25% quantile** | **75% quantile** | **Relative case increase %** | **Relative case increase 25% quantile** | **Relative case increase 75% quantile** |
|---|---|---|---|---|---|---|---|
| 20% | Two weeks | 41.5 | 25 | 62.25 | 28.71105 | 16.82474 | 44.98016 |
| 20% | One month | 36 | 21 | 55 | 25.00 | 14.65 | 38.68 |
| 20% | Two months | 20 | 6 | 40 | 13.90 | 3.83 | 29.55 |
| 60% | Two weeks | 84 | 58 | 127 | 56.50 | 38.75 | 90.82 |
| 60% | One month | 81 | 53.75 | 121 | 54.78 | 36.50 | 85.66 |
| 60% | Two months | 61 | 36.75 | 104.25 | 42.73 | 23.99 | 76.09 |
| 80% | Two weeks | 96 | 67.75 | 142 | 65.38 | 44.86 | 102.87 |
| 80% | One month | 93 | 62 | 141 | 63.72 | 42.41 | 99.35 |
| 80% | Two months | 77 | 47 | 128 | 54.10 | 31.87 | 89.39 |

### S9 $R_0$ calculation

$R_0$ was calculated using the expression for $R_0 = \beta[(1-\omega)(\frac{1}{\epsilon_1} + \frac{1}{\epsilon_2}) + \omega(\frac{1}{\alpha} + \frac{1}{\gamma_1} + ((1-\tau)\frac{1}{\gamma_2+d})]$ using the standard infection rate/output rates pattern (see Castillo-Garsow and Castillo-Chavez (2020) for more details).

### S10 Availability of codes and related data

Please refer to the link `https://github.com/PunyaAlahakoon/COVID_19_in_US_rural_counties.git`.
